# BrainAge moderates associations between Alzheimer’s disease biomarkers and cognitive decline: a meta-analysis across A4/LEARN, HABS and ADNI cohorts

**DOI:** 10.1101/2025.07.07.25331026

**Authors:** Jorge Garcia Condado, Hannah M Klinger, Colin Birkenbihl, Madison Cuppels, Annie Li, Iñigo Tellaetxe Elorriaga, Mabel Seto, Gillian T. Couglan, Michael J. Properzi, Dorene M. Rentz, Aaron P. Schultz, Asier Erramuzpe, Hyun-Sik Yang, Jasmeer Chhatwal, Keith A. Johnson, Brian C. Healy, Jesus M. Cortes, Reisa A. Sperling, Michael Donohue, Timothy J. Hohman, Ibai Diez, Rachel F. Buckley, Alzheimer’s Disease Neuroimaging Initiative

## Abstract

BrainAge delta, the difference between a person’s predicted brain age and their chronological age, is a promising marker of the accumulation of neurodegeneration that may increase vulnerability to Alzheimer’s disease (AD). In this study we use structural MRI regions vulnerable to AD, the hippocampus, the amygdala and cortical thickness measurements to build BrainAge models. We examined whether BrainAge delta moderates the relationship between AD biomarkers and longitudinal cognitive decline performing a meta-analysis across three cohorts: A4/LEARN, HABS and ADNI (2,279 cognitively unimpaired [CU]; 416 with mild cognitive impairment [MCI]). Higher BrainAge delta was linked to faster decline in CU (β = -0.13 [-0.21, - 0.06], p = 0.018, I^2^=1%) and more strongly in MCI (β = -0.31 [-0.30, -0.24], p < 1x10^-16^). BrainAge also interacted with Aβ-PET (β = -0.09 [-0.13, -0.05], p = 0.0054, I^2^=12%) and plasma pTau_217_ (β = -0.09 [-0.15, -0.03], p = 0.018, I^2^=0.1%), but not Tau-PET, to impact cognitive decline, where synergistically higher BrainAge delta and elevated AD markers resulted in faster cognitive decline. We next tested its utility for clinical trial enrichment. Sequential screening with pTau_217_ and BrainAge delta reduced required sample size for prevention trials by 77%, versus 61% using pTau_217_ alone. These findings support BrainAge delta as a marker of neurodegeneration and may serve as an enrichment tool for AD prevention trials.

## Background

BrainAge, a neuroimaging-based measure of brain aging, has been increasingly studied as a marker of accelerated brain aging and a potential biomarker of vulnerability to Alzheimer’s disease (AD)^1,2^. BrainAge is calculated by measuring the difference between a person’s predicted brain age, estimated from structural MRI, and their actual chronological age. This difference is referred to as the BrainAge delta. A positive BrainAge delta reflects accelerated or atypical brain aging. While established AD neuroimaging and biofluid biomarkers of amyloid-β (Aβ), Tau, and neurodegeneration (collectively forming the ATN framework)^3^, are the gold standard of AD pathological staging, their interaction with BrainAge in moderating cognitive trajectories remain incompletely characterized. This knowledge gap is particularly significant considering mounting evidence that AD pathology may accelerate brain aging processes, potentially offering a mechanistic link between conventional biomarkers, such as Aβ and Tau tangles, and cognitive decline observed in clinical settings^4,5^.

Previous cross-sectional studies have demonstrated associations between BrainAge delta and key AD biomarkers. Higher BrainAge delta is positively associated with greater Aβ-PET and Tau-PET burden^6^ at the cross-section and is also associated with *APOE*ε4 status^7^. When stratified by clinical diagnosis, these associations are primarily observed in patients with mild cognitive impairment (MCI) and AD dementia^6–8^. Higher BrainAge delta is also correlated with lower cognitive performance^9,10^. The importance of BrainAge delta to moderate the relationship between cross-sectional AD biomarkers and longitudinal cognitive decline remains poorly understood. Our overarching aim was to examine the extent to which baseline BrainAge delta informs rates of longitudinal cognitive decline in cognitively unimpaired (CU) and MCI individuals in the context of AD biomarkers using meta-analytic approaches across three cohorts. We also explored the potential application of BrainAge delta to enrich screening protocols for AD clinical trials by evaluating its use alongside plasma pTau_217_, a widely used early marker of elevated Aβ-PET burden and now increasingly used to screen for participants in early prevention trials such as AHEAD 3-45^11^, for participant selection^12^.

## Results

### Relationship between cross-sectional BrainAge delta and AD biomarkers

We analyzed data from 2279 cognitively unimpaired individuals from A4/LEARN (N=1635), HABS (N=299), and ADNI (N=345), as well as 416 individuals with MCI from ADNI. Demographic characteristics, including age, sex, education (in years), *APOE*ε4 status, cognitive performance, and biomarker measurements (Aβ-PET, pTau_217_, and Tau-PET), are summarized in **Table 1**. BrainAge models were trained using neuroimaging features from Aβ− and *APOE*ε4 non-carriers and then applied to other participants to estimate BrainAge delta for each participant (see **Figure 1**). Models were trained separately for each cohort.

**Figure 1.**
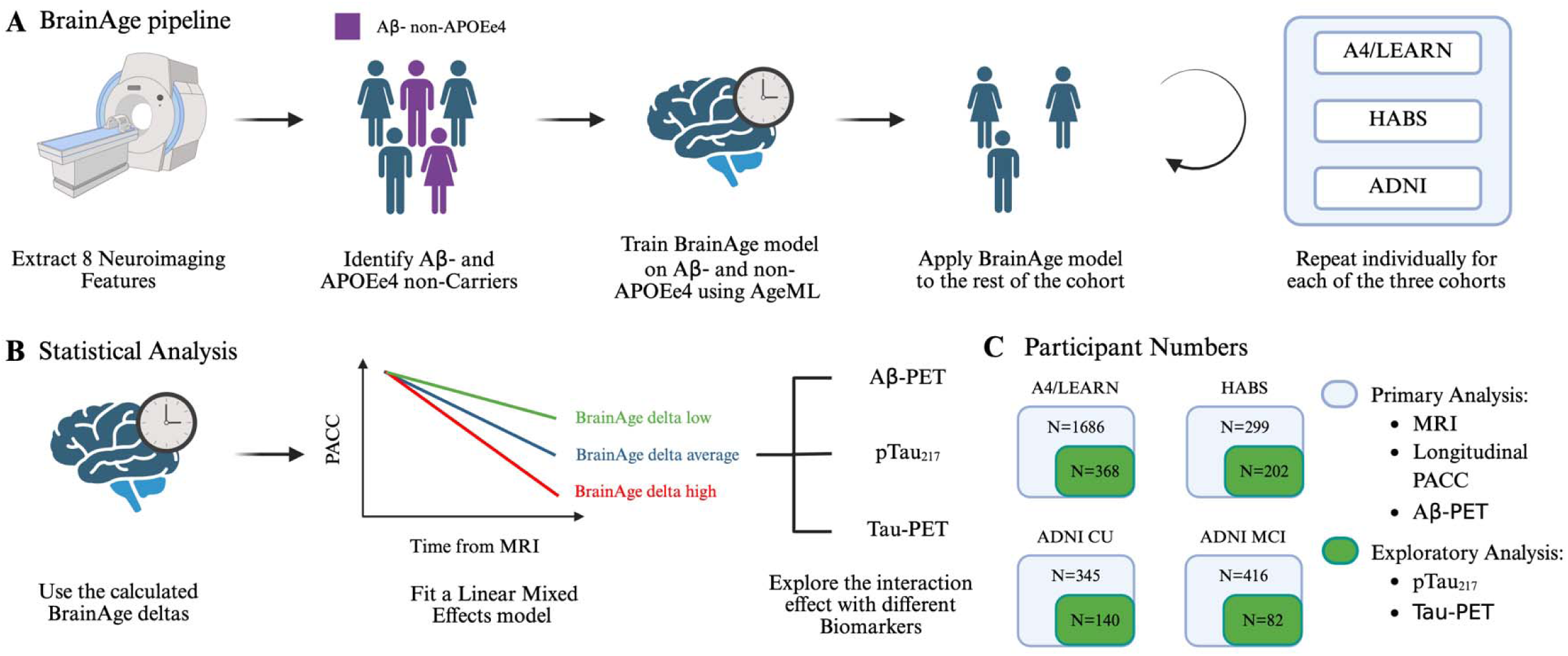
Overview of the study design to investigate BrainAge delta as a biomarker in Alzheimer’s disease. **(A)** BrainAge models were trained using neuroimaging features from Aβ− and *APOE*ε4 non-carriers and then applied to other participants to estimate BrainAge delta. Models were trained separately for each cohort. In ADNI the BrainAge model was trained on cognitively unimpaired participants and then applied to mild cognitive participants. **(B)** BrainAge deltas were used in a linear mixed-effects model to evaluate their interaction with longitudinal cognitive decline (PACC) and different biomarkers (Aβ-PET, pTau_217_, Tau-PET). **(C)** There was a primary analysis carried out with all participants that had an MRI scan, longitudinal PACC measurements and an Aβ-PET scan. An exploratory analysis was carried out with participant which had all previous datapoints and measurements of pTau_217_ and Tau-PET. Each square shows the number of participants in each cohort and analysis.

**Table 1.**
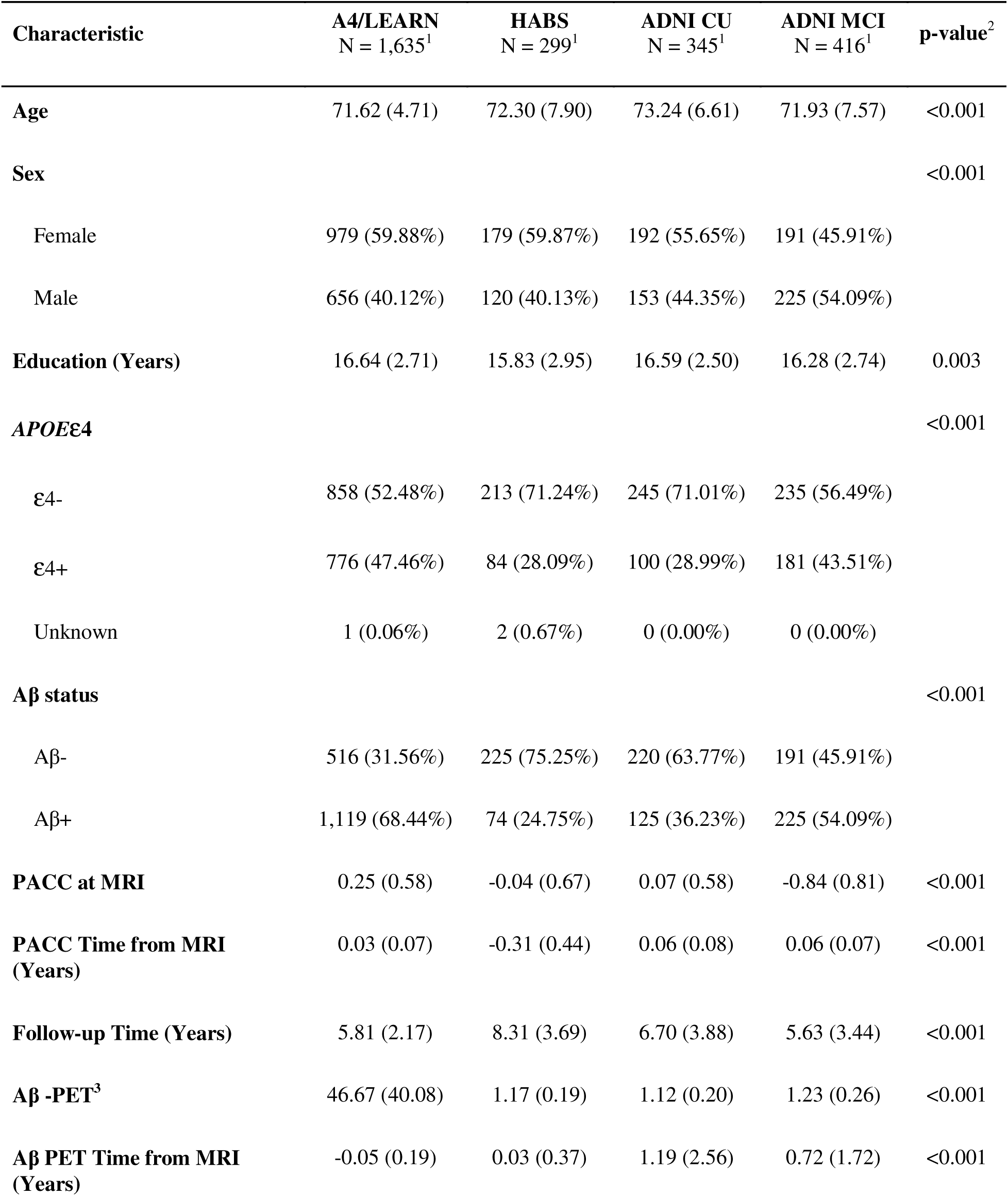
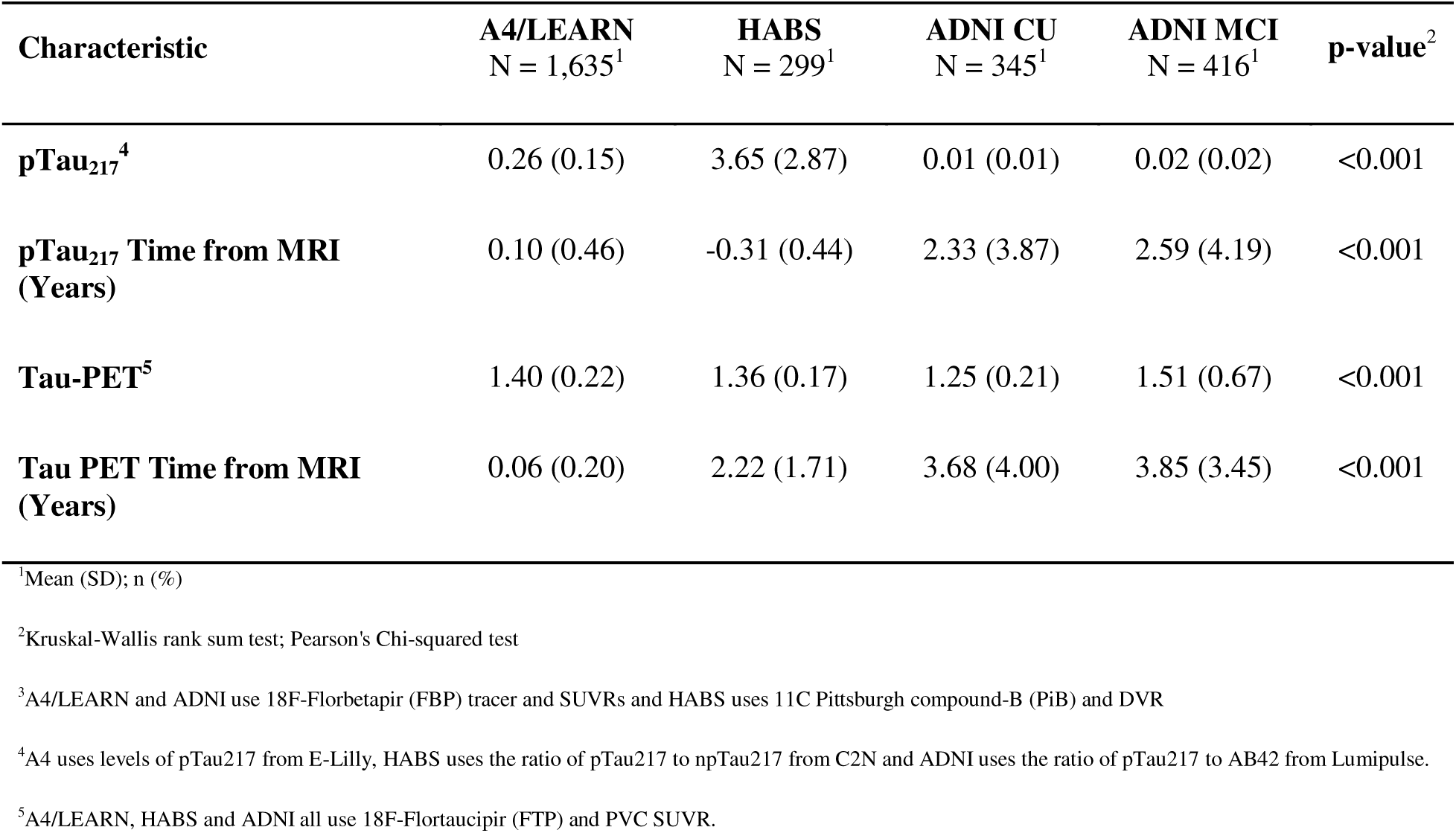
Sample demographic.

| Characteristic | A4/LEARN<br>N = 1,635<sup>1</sup> | HABS<br>N = 299<sup>1</sup> | ADNI CU<br>N = 345<sup>1</sup> | ADNI MCI<br>N = 416<sup>1</sup> | p-value<sup>2</sup> |
| --- | --- | --- | --- | --- | --- |
| **Age** | 71.62 (4.71) | 72.30 (7.90) | 73.24 (6.61) | 71.93 (7.57) | <0.001 |
| **Sex** |  |  |  |  | <0.001 |
| Female | 979 (59.88%) | 179 (59.87%) | 192 (55.65%) | 191 (45.91%) |  |
| Male | 656 (40.12%) | 120 (40.13%) | 153 (44.35%) | 225 (54.09%) |  |
| **Education (Years)** | 16.64 (2.71) | 15.83 (2.95) | 16.59 (2.50) | 16.28 (2.74) | 0.003 |
| **APOEε4** |  |  |  |  | <0.001 |
| ε4- | 858 (52.48%) | 213 (71.24%) | 245 (71.01%) | 235 (56.49%) |  |
| ε4+ | 776 (47.46%) | 84 (28.09%) | 100 (28.99%) | 181 (43.51%) |  |
| Unknown | 1 (0.06%) | 2 (0.67%) | 0 (0.00%) | 0 (0.00%) |  |
| **Aβ status** |  |  |  |  | <0.001 |
| Aβ- | 516 (31.56%) | 225 (75.25%) | 220 (63.77%) | 191 (45.91%) |  |
| Aβ+ | 1,119 (68.44%) | 74 (24.75%) | 125 (36.23%) | 225 (54.09%) |  |
| **PACC at MRI** | 0.25 (0.58) | -0.04 (0.67) | 0.07 (0.58) | -0.84 (0.81) | <0.001 |
| **PACC Time from MRI (Years)** | 0.03 (0.07) | -0.31 (0.44) | 0.06 (0.08) | 0.06 (0.07) | <0.001 |
| **Follow-up Time (Years)** | 5.81 (2.17) | 8.31 (3.69) | 6.70 (3.88) | 5.63 (3.44) | <0.001 |
| **Aβ -PET<sup>3</sup>** | 46.67 (40.08) | 1.17 (0.19) | 1.12 (0.20) | 1.23 (0.26) | <0.001 |
| **Aβ PET Time from MRI (Years)** | -0.05 (0.19) | 0.03 (0.37) | 1.19 (2.56) | 0.72 (1.72) | <0.001 |
| **pTau<sub>217</sub>**<sup>4</sup> | 0.26 (0.15) | 3.65 (2.87) | 0.01 (0.01) | 0.02 (0.02) | <0.001 |
| **pTau<sub>217</sub> Time from MRI (Years)** | 0.10 (0.46) | -0.31 (0.44) | 2.33 (3.87) | 2.59 (4.19) | <0.001 |
| **Tau-PET**<sup>5</sup> | 1.40 (0.22) | 1.36 (0.17) | 1.25 (0.21) | 1.51 (0.67) | <0.001 |
| **Tau PET Time from MRI (Years)** | 0.06 (0.20) | 2.22 (1.71) | 3.68 (4.00) | 3.85 (3.45) | <0.001 |

<sup>1</sup>Mean (SD); n (%)
<sup>2</sup>Kruskal-Wallis rank sum test; Pearson's Chi-squared test
<sup>3</sup>A4/LEARN and ADNI use 18F-Florbetapir (FBP) tracer and SUVRs and HABS uses 11C Pittsburgh compound-B (PiB) and DVR
<sup>4</sup>A4 uses levels of pTau217 from E-Lilly, HABS uses the ratio of pTau217 to npTau217 from C2N and ADNI uses the ratio of pTau217 to AB42 from Lumipulse.
<sup>5</sup>A4/LEARN, HABS and ADNI all use 18F-Flortaucipir (FTP) and PVC SUVR.

We investigated whether BrainAge delta is associated with *APOE*ε4 carriership, sex, PACC, Aβ-PET burden, pTau_217_ levels and Tau-PET burden in a meta-analysis of all CU cohorts and separately the ADNI MCI cohort. BrainAge delta was not a significantly associated with any of the variables in a meta-analysis of the CU cohorts (see **Figure 2**). On the other hand, higher BrainAge delta in the ADNI MCI cohort was associated with *APOE*ε4 carriership (β = 0.50 [0.29, 0.71], p < 1x10^-5^), Females (β = 0.29 [0.09, 0.49], p = 0.0036), lower PACC (β = -0.24 [-0.31, -0.15], p < 1x10^-9^), higher Aβ-PET (β = 0.31 [0.21, 0.41], p < 1x10^-8^), higher pTau_217_ levels (β = 0.29 [0.10, 0.47], p = 0.0032) and higher Tau-PET burden (β = 0.35 [0.14, 0.57], p = 0.0024). BrainAge deltas was also higher in ADNI MCI relative to ADNI CU (β = 0.54 [0.38, 0.70], p < 1x10^-10^). However, we saw that in A4/LEARN and HABS higher BrainAge delta was significantly correlated with higher Aβ-PET burden, higher pTau_217_ levels and higher Tau-PET burden (see **Supplementary Figure S1**).

**Figure 2.**
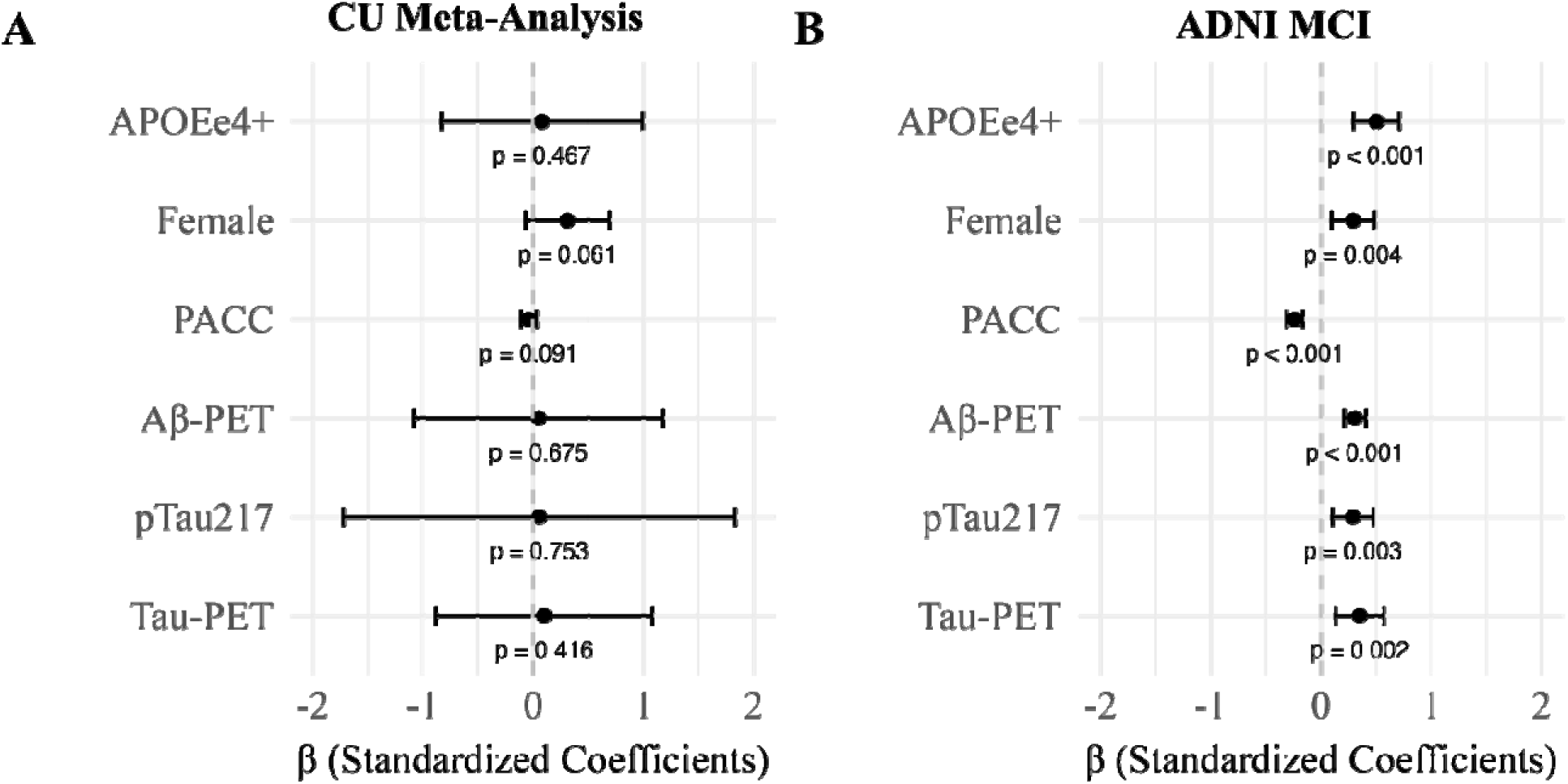
Standardized coefficients (β) representing the association between BrainAge delta and various biological and demographic factors in cognitively unimpaired (CU) individuals and ADNI MCI participants. (A) Meta-analysis of all CU cohorts **(B)** Analysis on ADNI MCI cohort.

### Influence of BrainAge delta on longitudinal cognitive decline

To quantify the effect of BrainAge delta on longitudinal cognitive decline, we conducted a meta-analysis across all cohorts and only the CU participants by building linear mixed effects models for each cohort (see **Figure 1)**. First, we compared linear vs quadratic models to model longitudinal cognitive decline, and we found quadratic models were a better fit and so pursued these in our subsequent analyses (see **Supplementary Table S2**). In a meta-analysis involving all cohorts we observed a significant association between higher BrainAge delta and faster cognitive decline (β = -0.17 [-0.34, -0.01], p = 0.046, I^2^=89%) (see **Figure 3** and **Supplementary Figure S2**). There is substantial heterogeneity most likely because the CU cohorts and MCI cohorts experience different effect sizes. When looking at a meta-analysis consisting of only CU participants, higher BrainAge delta was still associated with faster cognitive decline (β = -0.13 [-0.21, -0.06], p = 0.018, I^2^=1%) and the result of the meta-analysis is more homogenous. The effect was strongest in ADNI MCI participants (β = -0.31 [-0.39, - 0.24], p < 1x10^-16^). All results remained significant when covarying for age at MRI, sex, education, *APOE*ε4, and Aβ status (see **Supplementary Table S2**). In A4/LEARN, results remained significant when looking only at the A4 Placebo group.

**Figure 3.**
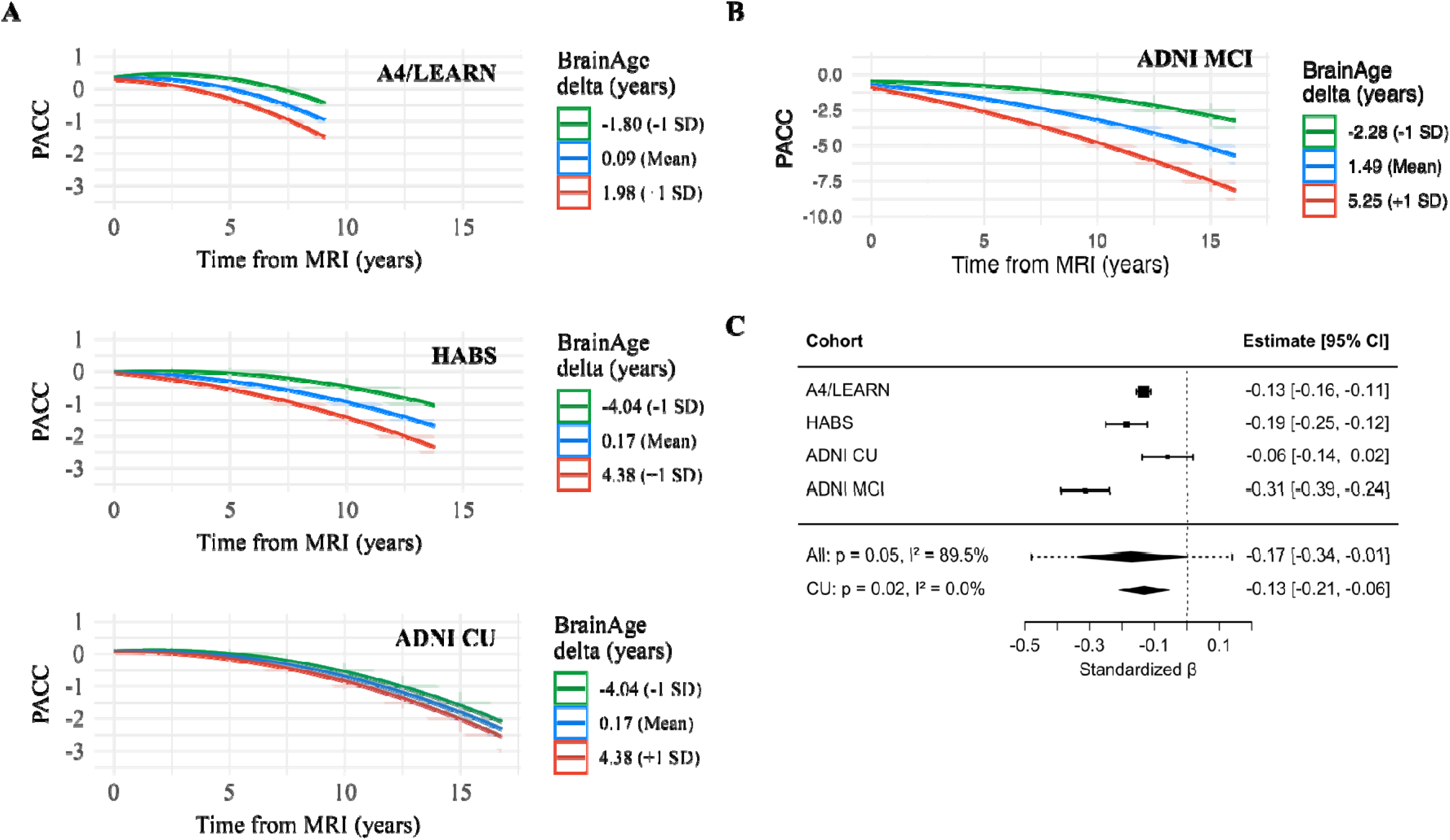
Association between BrainAge delta and longitudinal PACC trajectories. **(A)** Linear mixed effects models of longitudinal PACC trajectories stratified by BrainAge delta in cognitively unimpaired individuals from the A4/LEARN, HABS, and ADNI CU cohorts. The green line represents –1 SD BrainAge delta, the blue line represents the mean, and the red line represents +1 SD. **(B)** Same as (A), but for individuals with mild cognitive impairment (ADNI MCI cohort), who show more rapid cognitive decline. Note the different y-axis scale. **(C)** Forest plot summarizing the interaction coefficients between BrainAge delta and time on PACC across cohorts and meta-analysis run on CU cohorts and all cohorts. Error bars indicate 95% confidence intervals.

### Moderating effect of BrainAge delta and AD biomarkers on longitudinal cognitive decline

We examined whether baseline BrainAge delta moderates the relationship between AD biomarkers (Aβ-PET, pTau_217_ and Tau-PET) and longitudinal cognitive decline performing a meta-analysis across all cohorts. We found the interaction between Aβ-PET burden and BrainAge on longitudinal cognitive decline to be significant (β = -0.09 [-0.13, -0.05], p = 0.0054, I^2^=12%), with higher BrainAge delta and higher Aβ-PET burden synergistically associating with faster cognitive decline (see **Figure 4A**, with a visualization in A4/LEARN in **Figure 4B** and other cohorts in **Supplementary Figure S3**). Results remained largely significant when adjusting for age at MRI, sex, education, *APOE*ε4, and restricting to participants with Aβ-PET scans within one year of the MRI. Higher BrainAge delta and higher pTau_217_ levels were also synergistically associated with cognitive decline (β = -0.09 [-0.15, -0.03], p = 0.018, I^2^=0.1%); **Figure 4C** with a visualization in A4/LEARN in **Figure 4D**). Sensitivity analyses largely aligned with these effects. We did not find a significant interaction between Tau-PET burden and BrainAge on longitudinal cognitive decline (β = -0.09 [-0.22, 0.05], p = 0.14, I^2^=74%), and heterogeneity was high in this pooled estimate (as evidenced in I^2^). We did note that these effects did appear in the A4/LEARN and HABS cohorts (**Figure 4E** with a visualization in A4/LEARN in **Figure 4F**). Individual interaction terms between time from MRI and BrainAge delta or the different biomarkers can be found in **Supplementary Table S3**, while all sensitivity analyses are presented in **Supplementary Table S4** and **S5**.

**Figure 4.**
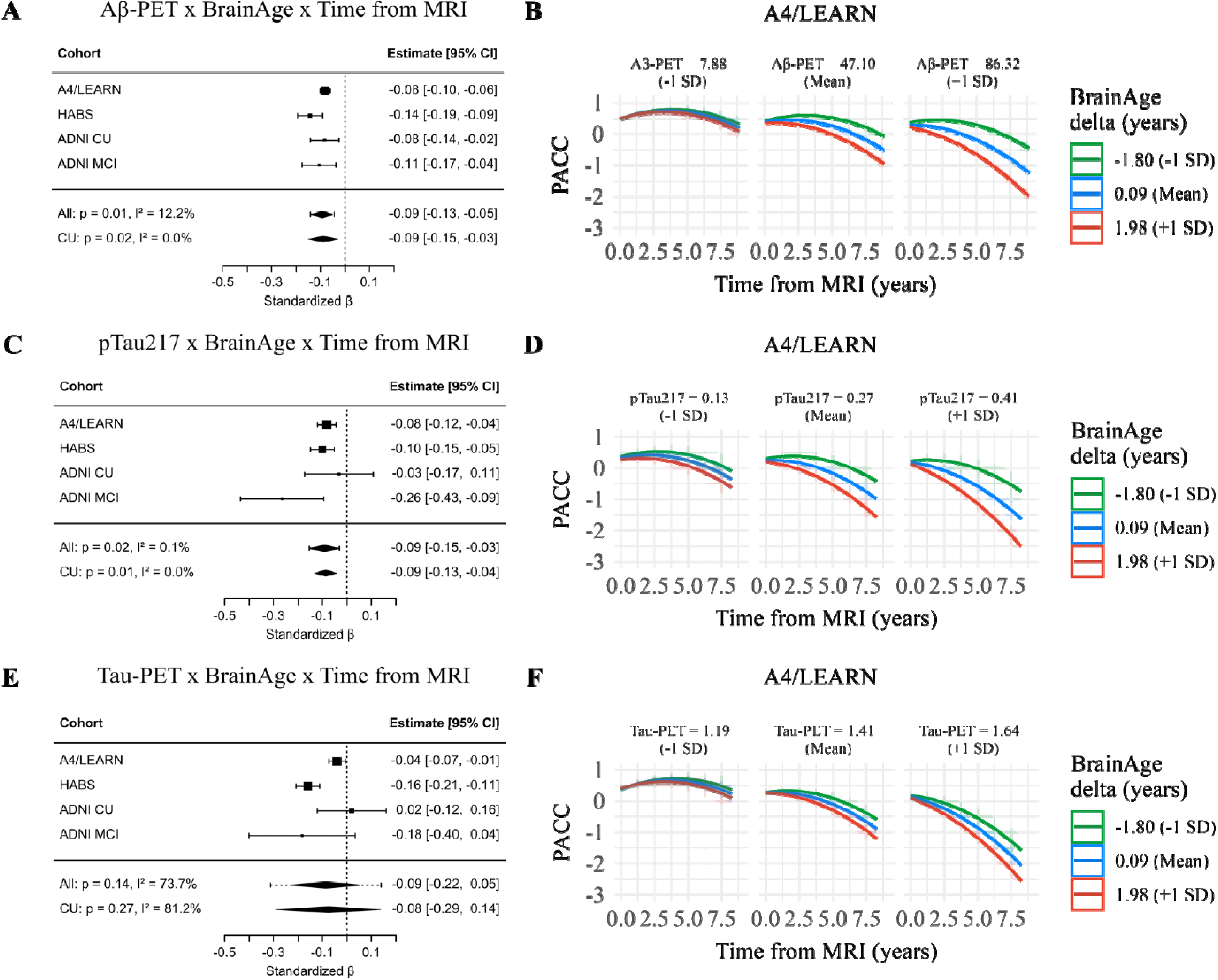
Interaction between BrainAge delta and Alzheimer’s disease biomarkers in moderating longitudinal cognitive decline. **(A)** Forest plot summarizing the interaction coefficients between BrainAge delta, Aβ-PET and time on PACC across cohorts. **(B)** Interaction between BrainAge delta and Aβ-PET (centiloids) in A4/LEARN Cohort stratified by BrainAge delta (–1 SD: green, mean: blue, +1 SD: red) and biomarker levels (columns: –1 SD, mean, +1 SD for Aβ-PET). **(C)** Forest plot summarizing the interaction coefficients between BrainAge delta, pTau_217_ and time on PACC across cohorts. **(D)** Interaction between BrainAge delta and pTau_217_ in A4/LEARN Cohort. **(E)** Forest plot summarizing the interaction coefficients between BrainAge delta, Tau-PET and time on PACC across cohorts. **(F)** Interaction between BrainAge delta and Tau-PET in A4/LEARN Cohort.

### Clinical trial enhancement

We explored how screening participants for BrainAge delta could reduce the number of participants required for an enriched AD trial focused on MCI participants. Screening participants with BrainAge delta above the median could reduce the required sample size by 40%, from 1,8889 participants using the original Clarity AD trial criteria^13^ to 1,139 participants, while maintaining 90% statistical power. To recruit this enriched cohort, approximately 7,533 individuals would need to be screened - an increase of 1,259 additional screens compared to the 6274 individuals screened in the conventional trial. This increased screening requirement would raise screening-related costs by approximately $9.6 million. The 24-month treatment costs for the lecanemab group (50% of participants) would be approximately $50.0 million in the conventional trial and $30.2 million in the enrichment trial. Overall, the estimated total cost of the conventional trial would be approximately $93.7 million, while the enrichment trial would cost approximately $83.4 million - yielding an overall cost savings of 11% (∼$10.3 million), despite the higher screening costs (see **Table 2)**. The linear mixed effects models used to estimate sample sizes for the conventional trial and enriched trial can be seen in **Supplementary Figure S4**.

**Table 2.** Comparing the estimated monetary expenses of a conventional trial to an enrichment trial, assuming screening for BrainAge delta above the median and using ADNI MCI Aβ+ subjects and an estimated trail duration of 2 years.

|  | Conventional Trial | Enrichment Trial | Difference |
| --- | --- | --- | --- |
| **Screened patients** | 6274 | 7533 | 1259 |
| **Screen pass before MRI** | 1889 | 2278 | 389 |
| **Recruited patients** | 1889 | 1139 | -750 |
| **Total screen cost** | \$43,653,878 | \$53,235,903 | \$9,582,02 |
| **Total treatment cost** | \$50,049,184 | \$30,192,022 | -\$19,857,162 |
| **Total costs** | \$93,703,061 | \$83,427,925 | -\$10,275,136 |

Next, we tested whether and how a two-step sequential approach (that is, plasma pTau_217_ followed by an MRI for BrainAge screening) could reduce the number of participants needed for a preclinical AD trial using longitudinal changes in cognitive function as the primary outcome. Substantial sample size reductions can already be achieved in the first step, that is, using only plasma pTau_217_. When using data from the A4/LEARN trial, PACC as the outcome measure, assuming 80% power and α = 0.05 in a 4-year clinical trial with repeated testing every 6 months, selecting participants with plasma pTau_217_ levels in quartiles 2–4 (i.e., Q2–Q4, which excludes the lowest 25% of plasma pTau_217_) would result in a 42% (26%-54%) reduction in the number of required participants compared to including the entire study population (**Figure 5**). Selecting participants in plasma pTau_217_ Q3–Q4 further reduced the required sample size by 61% (48%-70%). Screening for BrainAge delta further reduced the required sample size needed in a clinical trial. For example, selecting for participants based on plasma pTau_217 in_ Q3-Q4 and BrainAge in Q3-Q4 together would reduce the sample size by 77% (68%-83%) (**Figure 5**). Repeating the same set of analyses but now restricted to Aβ-positive CU individuals (i.e., excluding participants from the LEARN group) showed that similarly large sample size reductions can be achieved in this population (**Supplementary Figure S5**).

**Figure 5.**
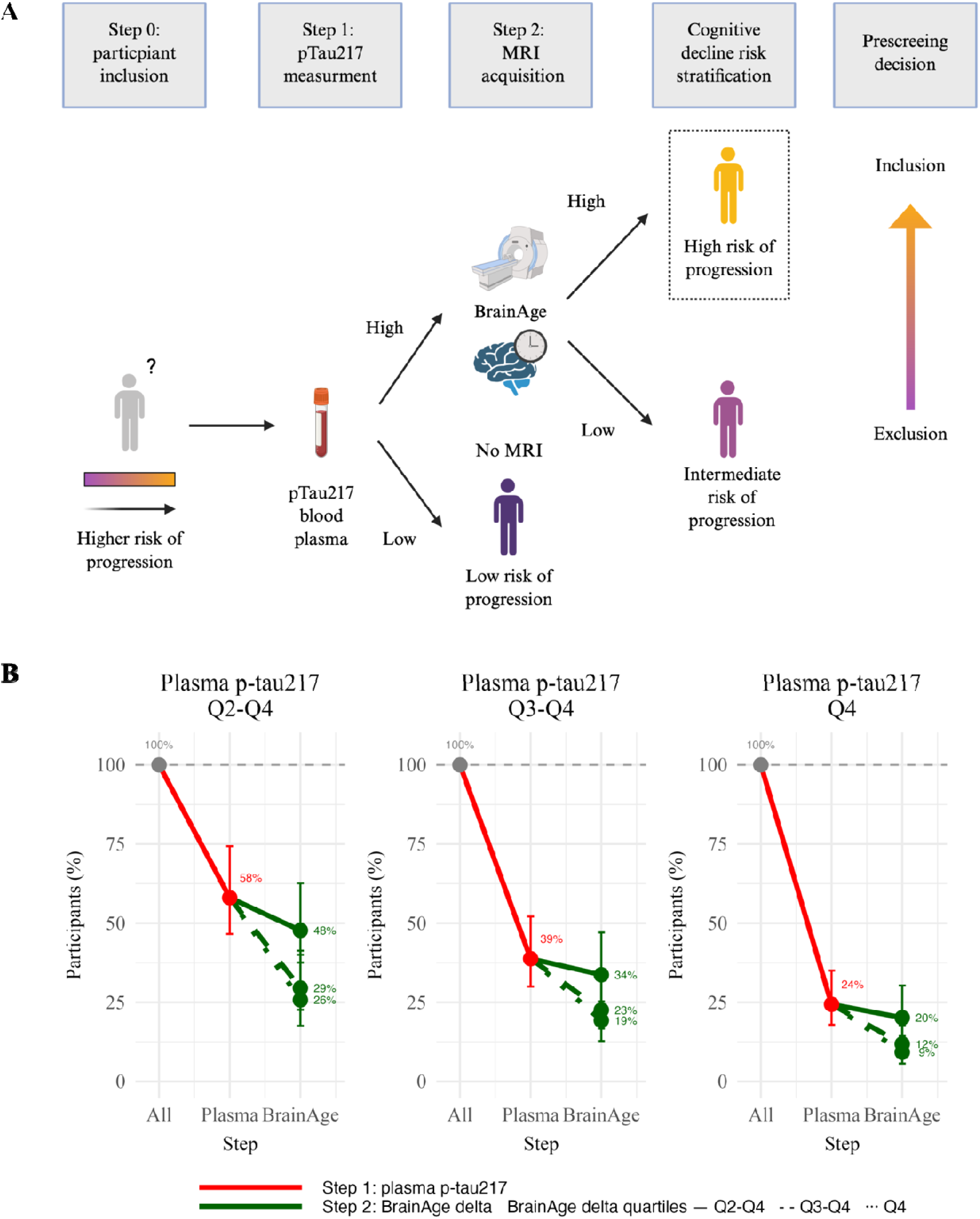
Two step recruitment strategy for Preclinical Alzheimer’s Clinical Trial. **(A)** Conceptual framework of a sequential two-step recruitment strategy of a clinical trial in preclinical AD using a cognitive endpoint. We followed procedures similar to those by Ossenkopple et al. 2025^18^ **(B)** The obtained sample size reduction using sample selection based on different percentiles (75^th^ [Q2-Q4], 50^th^ [Q3-Q4] and 25^th^ [Q4]) of baseline plasma pTau_217_ levels in step 1 followed by selection based on the same percentiles (75th, 50th and 25th) of the BrainAge measurement in step 2 with PACC as the primary endpoint. Error bars represent the 95% CI around the mean derived from linear effects models. Red lines represent step 1 with plasma p-tau217, and green lines represent step 2 with BrainAge. Different line styles represent different quartiles of BrainAge from those participants already selected from step 1. Calculations assume 80% power to detect a 30% change in PACC in a 4-year trial.

## Discussion

We found that cross-sectionally higher BrainAge delta was consistently associated with elevated AD biomarkers in MCI participants but not CU participants. Our findings were strongest in the ADNI MCI cohort where we found significant correlations with Aβ-PET burden, pTau_217_ and Tau-PET, as well as with cognitive performance. By contrast, the ADNI CU cohort, which had a lower prevalence of high Aβ, the patterns of effects were much weaker. Consistent with previous studies^6,8,14^, we argue that the strength of these cross-sectional associations appears to be linked with level of cognitive impairment. We also found that individuals with higher BrainAge delta experienced faster rates of cognitive decline, particularly in MCI, aligning with recent reports suggesting that BrainAge captures latent brain vulnerability associated with cognitive decline^15^. These patterns suggest that the utility of BrainAge delta as a marker of vulnerability to AD clinicopathology may be more pronounced in individuals already experiencing cognitive decline or with more established biomarker evidence of the disease.

We found BrainAge delta and Aβ-PET burden synergistically moderated cognitive decline across all four cohorts suggesting that elevated BrainAge delta potentially reflects the accumulation of neurodegenerative processes which increases the susceptibility to the effects of elevated Aβ pathology. We found these effects to be significant even in CU participants. BrainAge delta serves as a general biomarker of brain health by quantifying the degree of atrophy relative to normative aging. Thus, given these effect sizes are similar across the four cohorts, BrainAge may reflect a vulnerability to cognitive decline that is concurrent with the onset of Aβ pathology. In exploratory analyses with smaller sample sizes, we observed that BrainAge delta also interacted with plasma pTau_217_ levels, and less consistently with Tau-PET, to influence cognitive decline. The lack of consistency of the Tau-PET results may be due to the small sample sizes as Tau-PET was introduced later on in these studies. Previous studies have shown an association between BrainAge and glial reactivity and synaptic dysfunction in cerebrospinal fluid biomarkers^16^. Taken together, these findings suggest that pre-existing brain aging, as captured by BrainAge delta, may reflect vulnerability to cognitive decline in the presence of AD pathology, highlighting a potential role for age-estimated brain volume and thickness measures to reflect a latent quality of the emergence of Aβ or Tau.

We examined the possibility of BrainAge delta to enrich clinical trials by selecting individuals more likely to show measurable cognitive decline, increasing statistical power and reducing the total number of participants needed. We reported, both in clinical trials with MCI patients, as well as prevention trials involving CU individuals, significant cost reductions and a lower sample needed to treat by including this measure. This enrichment strategy could be particularly beneficial for trials targeting individuals with MCI. Notably, we found that BrainAge delta could further reduce sample size requirements even in preclinical AD prevention trials beyond the predictive utility of plasma pTau_217_ alone. Plasma pTau_217_ is fast becoming a critical element of screening processes for AD clinical trials^17,18^. Screening for pTau_217_ in our simulations based on A4/LEARN results in a similar sample size reduction to those results published by Ossenkopelle et al.^18^ using a different set of cohorts. Our findings suggest that BrainAge delta could serve as a valuable complementary measure to capture a quantifiable measure of brain health even after screening for pTau_217_. Sequential screening with MRI-based BrainAge estimation could be implemented after the cognitive and medical measures and blood test, but prior to the more expensive and invasive PET scan. This approach has the potential to reduce screen failure rates and overall trial costs by identifying individuals with evidence of accelerated brain aging who may be more likely to exhibit cognitive decline. Moreover, by potentially reducing the total number of participants required for a trial, this enrichment strategy could also minimize overall exposure to experimental treatments and the associated risk of adverse events.

While our findings demonstrate that selecting using the BrainAge delta substantially reduces sample sizes for AD clinical trials, selecting participants based on greater-than-expected neurodegeneration may paradoxically identify those least likely to respond to disease-modifying interventions targeting upstream pathology. This paradox was notably observed in recent anti-amyloid monoclonal antibody trials, such as CLARITY AD^13^ and TRAILBLAZER-ALZ-2^19^, where participants with lower amyloid and tau burden at baseline showed substantially larger treatment effect sizes, while those with more advanced neurodegeneration showed diminished treatment responses. Therefore, predictors of faster cognitive decline were fundamentally different from predictors of therapeutic response. This reflects a critical tension in AD prevention trial design: achieving sufficient cognitive decline to detect treatment effects within a reasonable timeframe requires selecting participants with established neurodegenerative changes, yet such markers of rapid decline may be associated with biological states less responsive to targeted interventions. Therefore, while BrainAge delta effectively enriches for cognitive decline, this enrichment does not guarantee enhanced therapeutic responsiveness, potentially necessitating larger treatment effect sizes to achieve statistical significance. Selecting for higher BrainAge delta might also increase the risk of adverse events such as ARIA (Amyloid-Related Imaging Abnormalities)^20^ as these are driven by more advanced pathology and we might be selecting for those types of participants. However, this also opens up the possibility to use the BrainAge delta for the opposite utility, to select those with lower vulnerability to neurodegeneration but with elevated AD biomarkers to optimize for treatment response.

A second important limitation concerns the biological specificity of BrainAge delta as a marker. Rather than solely reflecting AD-specific pathology, BrainAge delta captures multiple aging-related neurobiological processes that extend beyond Alzheimer’s pathology^21^. Notably, BrainAge is affected by cerebrovascular disease and cerebrovascular disease accelerates tau accumulation when combined with amyloid-beta and independently contributes to cognitive decline through direct effects on neuronal integrity, particularly in APOEε4 carriers^22^. This biological heterogeneity has critical implications for clinical trial interpretation: while selection for elevated BrainAge delta effectively selects for faster cognitive decline, it may enrich not only for AD-specific neurodegeneration but also for non-AD neurodegenerative processes. If a substantial portion of the enriched cohort experiences cognitive decline through amyloid-independent mechanisms (such as primarily vascular contributions), therapeutic responses to amyloid-directed agents would be attenuated, effectively reducing the observable treatment effect size. In this scenario, the theoretical sample size savings could be offset by diminished treatment efficacy in this more neurodegenerate population. However, BrainAge models specifically trained to capture only AD related neurodegeneration could help reduce this problem^22^. Clinical trials should therefore carefully consider whether enriching for neurodegeneration markers selects for participants with pathophysiological mechanisms aligned with the therapeutic target.

Several limitations should be considered when interpreting our findings. Specific assays used for pTau_217_ measurements and Aβ tracers varied across the different cohorts. Second, most participants in our study were non-Hispanic White, limiting the generalizability of our findings to more diverse populations. Additionally, the sample sizes that included plasma pTau_217_ and Tau-PET were relatively small, which may have limited the statistical power to detect more subtle effects. Heterogeneity in cohort sampling, inclusion criteria, and baseline participant characteristics across the three cohorts may have also influenced model performance and generalizability of the trial enrichment simulations, particularly regarding BrainAge delta thresholds and projected screen fail rates. Finally, further research is needed to standardize BrainAge estimation pipelines and establish robust cutoffs for clinical applications. This includes harmonizing model training procedures and gaining a clearer understanding of how BrainAge delta should be interpreted across various populations and stages of Alzheimer’s disease.

In conclusion, BrainAge delta is a biomarker of neurodegeneration and a biomarker of vulnerability to AD pathology. Cross-sectionally, higher BrainAge delta is associated with greater AD biomarker burden and lower cognition in MCI. Longitudinally, higher BrainAge delta is associated with faster cognitive decline, and in combination with elevated AD biomarkers, is synergistically linked to steeper decline in both CU and MCI participants. BrainAge delta may be of utility to enrich clinical trials to increase statistical power and reduce the total number of participants needed. Our findings support BrainAge delta as a quantifiable measure of vulnerability to neurodegeneration with potential to advance clinical translation in AD research.

## Methods

### Sample characteristics

2279 cognitively unimpaired (CU) individuals were sampled from Anti-Amyloid Treatment in Asymptomatic Alzheimer’s Disease trial and the companion Longitudinal Evaluation of Amyloid Risk and Neurodegeneration study ^23^ (A4/LEARN, N=1635), Harvard Aging Brain Study ^24^ (HABS, N=299) and Alzheimer’s Disease Neuroimaging Initiative ^25^ (ADNI, N=345). An additional 416 individuals with MCI were sampled from ADNI, with evaluations of MCI in ADNI described elsewhere^25,26^. We conducted the procedures for this study under the ethical guidelines stipulated by the Massachusetts General Brigham Human Research Committee, which is the relevant Institutional Review Board. The study was carried out by the guidelines of the Declaration of Helsinki. Written informed consent was obtained from all participants.

For all analyses, participants had to have an MRI scan and have completed at least two cognitive assessments with at least 6 months of follow-up. For analyses involving Aβ-PET, we used data from 2695 participants who met the aforementioned requirements and had at least one an Aβ-PET acquisition. The Aβ-PET scan acquired at the closest available time point to the first MRI scan was selected. For analyses involving plasma pTau_217_ and Tau-PET, we included all participants from the primary analysis who also had both a plasma pTau_217_ measurement and a Tau-PET scan (N=782), again we used the biomarker acquisition closest in time to the MRI. We split the analysis into these two samples due to the lower number of subjects who have pTau_217_ and Tau-PET scans and as Tau-PET was introduced later on in each of the studies. **Supplementary Table S1** provides demographic details separately for participants included in the primary and exploratory analyses.

### Imaging Measures

The first available T1-weighted MRI image of each participant was processed using FreeSurfer 7.4.1^27^ for all three cohorts: A4/LEARN, HABS and ADNI. 8 neuroimaging features were extracted, 2 volumetric measurements and 6 cortical thickness measurements. The 2 volumetric measurements were the bilateral hippocampus and amygdala volumes normalized with respect to the total intracranial volume. The hippocampus^28^ and amygdala^29^ were chosen as they are AD vulnerable regions affected early in preclinical AD. The 6 cortical thickness measurements were taken from the entorhinal, fusiform, inferior parietal, inferior temporal, middle temporal, and parahippocampal. Cortical thickness measurements were chosen as AD is reliably associated with a cortical signature of thinning^30^.

A4/LEARN and ADNI used the 18F-Florbetapir (FBP) tracer for Aβ-PET acquisition, whereas 11C Pittsburgh compound-B (PiB) was used in HABS. The PET acquisition parameters for each study are described elsewhere^24,31,32^. The processing pipeline for Aβ-PET data has been described elsewhere^24,31,32^. Aβ-PET data were treated as continuous, however, Aβ status, where referenced, was based on the published study-specific data cutoffs. We report Centiloid^33^ values for ease of comparison across cohorts^34–36^.

Tau-PET was performed using the 18F-Flortaucipir (FTP) tracer in all three cohorts. The study-specific FTP-PET acquisition parameters and preprocessing for each study are described elsewhere^37–39^. In all three cohorts, we used an SUVr with Partial Volume Correction (PVC) based on spreadsheet data available in each cohort. We then derived bilateral meta-temporal composites from an unweighted average of the amygdala, entorhinal, fusiform, inferior parietal, inferior temporal, middle temporal, and parahippocampal and referenced to cerebellar grey. These regions were chosen as they are regions affected early in preclinical AD^40^. SUVRs from the resulting composites were not harmonized across cohorts.

### Cognitive outcome measures

Cognition was measured using the Preclinical Alzheimer Cognitive Composite score (PACC)^41^. In HABS and ADNI, we used the PACC-5 composite, which includes a semantic processing test^42^. HABS includes the MMSE total score, Logical Memory Delayed Recall (Anna Thompson story), the Digit Symbol Substitution Test (WAIS-R; 90s), Category Fluency (animals, vegetables, fruits), and the Free and Cued Selective Reminding Test (FCSRT)^43^. The ADNI PACC is comprised of the MMSE total score, Logical Memory Delayed Recall (Anna Thompson story), Trails B time, Category Fluency (Animals), and ADAS-Cog Delayed Word Recall^44^. A4/LEARN, however does not include a semantic processing test, and so only the standard version of the PACC was used^41^. A4/LEARN uses the MMSE total score, Logical Memory Delayed Recall (Robert Miller story), the Digit Symbol Substitution Test (WAIS-R; 90 s), and the Free and Cued Selective Reminding Test (FCSRT)^45^. For A4/LEARN, alternative versions of the component subtests were administered across visits to minimize practice effects, and a time-varying covariate indicating the PACC version was incorporated into the analyses. Longitudinal measurements of PACC had a mean follow up time of 5.81 [0.53-9.17] years for A4/LEARN, 8.31 [0.86-13.86] years for HABS and 6.70 [0.55-16.90] years ADNI CU and 5.63 [0.59-16.11] years ADNI MCI. PACC measurements were harmonized across cohorts for ease of comparison across cohorts based on Hampton et al.^44^.

### APOE genotyping

Direct genotyping of *APOE* was performed in each study through blood sample collection. *APOE*ε4 carriership was treated as a binary variable, defined as carrying at least one copy of the ε4 allele.

### Blood biomarkers

Measurements of levels of pTau_217_ closest to the first MRI were used for each of the cohorts. In A4/LEARN, pTau_217_ levels were measured by Eli Lilly and Company using an electrochemiluminescent immunoassay, with sample preparation automated by the Tecan Fluent workstation and detection performed on the MSD Sector S Imager 600MM^46^. In HABS, plasma samples were assayed with C2N Diagnostics (mass spectrometry) to calculate a pTau_217_/npTau_217_ ratio^47^. In ADNI, the ratio of pTau_217_ to Aβ_42_ were measured on the Fujirebio Lumipulse G1200 automated immunoassay platform^48^. We did not harmonize plasma measures due to the lack of established harmonization procedures.

### BrainAge modelling

For each cohort, we built separate BrainAge models using AgeML^49^, an Open-Source tool for age modelling. Methodological and technical details of the age modelling process are described in AgeML^49^. To train the BrainAge model, we used the 8 neuroimaging features described in Section Imaging Measures. BrainAge models for each cohort are trained on CU individuals who are *APOE*ε4 non-carriers and have low Aβ burden (based on established cut-offs) to avoid biased estimates^50^. For each participant, we calculated their BrainAge with respect to their age at their respective baseline MRI. A linear regressor was used as the base model. We used a 5-fold cross validation strategy for training. The model trained on the *APOE*ε4 non-carriers and Aβ negative sample, was then applied to the remaining participants of the cohort. The BrainAge delta was calculated as the difference between the age predicted by the model after age bias correction and the respective participant’s chronological age.

### Statistical Analysis

All statistical analyses were performed in R version 4.4.2 (The R Foundation, Vienna, Austria). For ADNI, analyses were carried out separately for CU and MCI participants. First, we examined whether BrainAge can predict *APOE*ε4 status and sex using a logistic regressor trained separately for each cohort. We then explored whether cross-sectional BrainAge delta can predict the closest-in-time measurement of Aβ-PET burden, pTau_217_ and Tau-PET, respectively, using a general linear model trained separately for each cohort. Following the extraction of estimates of interest, a meta-analysis was conducted on all CU cohorts to generate standardized coefficients (β) using the *metafor* package in R. These were compared to the coefficients from the ADNI MCI cohort.

We used a series of linear mixed-effects models including random intercepts and slopes to examine the relationship between BrainAge delta and longitudinal cognitive decline separately for each cohort. Given that curvilinear models tend to fit longitudinal cognitive decline in these studies^51,52^, we examine linear and quadratic models, using ANOVA fit indices to determine best fit. We also explored the moderating effects of BrainAge delta with baseline Aβ-PET burden, pTau_217_ and Tau-PET burden on cognitive decline. Time was measured as the years between the date of PACC measurement and the first MRI scan of each participant. For the A4/LEARN model, we adjusted for cohort (A4 Treated vs A4 Placebo vs LEARN), PACC version, and cumulative dose to align as closely as possible with the models that were applied in the original clinical trial^45^. The primary models were:

1. Longitudinal PACC ∼ Time^2^* + Brain Age delta × Time
2. Longitudinal PACC ∼ Time^2^ + Brain Age delta × Aβ-PET × Time
3. Longitudinal PACC ∼ Time^2^ + Brain Age delta × Tau-PET × Time
4. Longitudinal PACC ∼ Time^2^ + Brain Age delta × pTau_217_ × Time

*We tested both linear only models as well as models with a quadratic term added to examine fit.

We also carried out sensitivity analyses that added sex, baseline age, *APOE*ε4, years of education and Aβ status as covariates in the model. For the A4/LEARN model we re-ran analyses using only the placebo and LEARN groups to ensure that there were no extraneous effects of treatment.

Following the extraction of estimates of interest, a meta-analysis was conducted on all model outputs to generate the global fixed effect (β) using the *metafor* package in R. A meta-analysis was run on all four cohorts and separately for the CU cohorts (A4/LEARN, HABS and ADNI CU).

### Clinical trial enrichment for Alzheimer’s Disease trials

To assess the effect of screening for BrainAge delta to enrich standard AD clinical trials with individuals who are most likely to experience cognitive decline, we simulated a randomized controlled trial (RCT) for a potential disease-modifying drug based on the eligibility criteria and study design for ‘Clarity AD’ trial for lecanemab^13^ To conduct these simulations, we followed the same methodology from Birkenbihl and colleagues^53^. We first selected 225 Aβ positive ADNI MCI participants who matched the eligibility criteria of the Clarity AD trial^13,54^. We set up the model to mirror the original design of the ‘Clarity AD’ trial of measuring Clinical Dementia Rating-Sum of Boxes (CDR)^55^ every 6 months for a total of 24 months of observation time^13^. We also selected 90% power and an alpha level (Type I error) of 0.05 and a treatment effect of 27% reduction in cognitive decline as measured by CDR Sum of Boxes^13^. For simplification, we used equally sized control and treatment groups without different treatment dosage arms. Sample size was calculated using a linear mixed-effects model, and assuming a linear increase of CDR over time. Calculations were based on a method from Edland et al.^56^ and implemented using the R package *longpower*^57^. Sample size calculations were performed using: (1) only participants with high Aβ and (2) only participants with high Aβ and BrainAge delta above the median. The linear mixed effects models were only trained with 24 months of data to approximate the duration of a clinical trial.

We estimated the impact of reducing the trial sample size in terms of monetary expenses using the Clarity AD trial as guidance^13^ and methodologies similar to previous studies^58^. We ignored patient dropout in our estimations. We estimated annual treatment costs of lecanemab at $26,500 per patient^59^. As exact information about the costs of patient screening in the Clarity AD trial was missing, we assumed the same costs that were previously estimated for the aducanumab trial with $6957 per screened patient^60^. For the trials involving calculating the BrainAge delta during screening we added to the cost an extra $353 per patient for the MRI scan^60^.

To assess the effect of screening for BrainAge delta and pTau_217_ to enrich new preclinical AD clinical trials with individuals who are most likely to experience cognitive decline, we simulated a randomized controlled trial (RCT) for a potential disease-modifying drug. We used all participant from the A4/LEARN dataset with pTau_217_ measurements and followed procedures similar to those by Ossenkople et al^18^. To derive optimal sample size reduction for a clinical trial in the two-step approach, we generated a data-driven estimate of the complementary value of BrainAge delta and plasma pTau_217_ when implementing a sequential two-step approach (that is, plasma pTau_217_ first, followed by BrainAge delta). We calculated the obtained sample size reduction when assuming 80% power to detect a 30% change in cognitive change (PACC) in a 4-year clinical trial (with repeated testing every 6 months). Sample size was then defined by using different percentiles (75th, 50th and 25th) of the participants’ baseline plasma pTau_217_ levels using a method from Edland et al.^56^ and the *lmmpower* function in the *longpower* package^57^. The approach was repeated, selecting the 75th, 50th and 25th percentiles of the new participants BrainAge delta.

## Data Availability

Data for A4/LEARN and HABS is available through Synapse for registered researchers. Data for ADNI is available through LONI for registered researchers. All code is available through Github.

https://github.com/JGarciaCondado/AgingBrainStudy

https://www.synapse.org/Synapse:syn61250768/wiki/628717

https://www.synapse.org/Synapse:syn53910452/wiki/626438

https://adni.loni.usc.edu/data-samples/adni-data/

## Acknowledgments

The research that led to this publication was conducted with the support of a US-Spain Fulbright grant. The project that gave rise to these results received the support of a fellowship from “la Caixa” Foundation (ID 100010434) to J.G.C. The fellowship code is LCF/BQ/DI21/11860030. J.M.C acknowledges financial support from the Health Department of the Basque Country (grants 202211103 and 2023111002). M.S. is supported by a research fellowship from the Alzheimer’s Association (24AARF-1201281). G.T.C. is funded on the National Institute on Aging (K99 AG083063), as well as a research fellowship from the Alzheimer’s Association (AARF-23-1151259). A.E. is supported by the Spanish Ministry of Science and Innovation, grant RYC2021-032390-I and the Basque Country’s Department of Economic Development and Infrastructure (Elkartek Program) under Grant KK-2024/00028. J.M.C., A.E. and I.D. are supported by Ikerbasque: The Basque Foundation for Science. J.M.C and A.E. are supported by the Spanish Ministry of Science under Grant PID2023-148008OB-I00. J.M.C. acknowledges financial support from the Basque Ministry of Health (grants 2023111002 & 2022111031). R.F.B. is supported by the National Institute on Aging (R01AG079142 and DP2 AG082342). I.D. was supported by Alzheimer’s Association (AARF-23-1145358) and Spanish Ministry of Science (RYC2022-035429-I and PID2023-150633OA-I00).

The A4 study is funded by a public-private-philanthropic partnership, including funding from the National Institutes of Health-National Institute on Aging (U19AG010483; R01AG063689), Eli Lilly and Company, Alzheimer’s Association, Accelerating Medicines Partnership, GHR Foundation, an anonymous foundation and additional private donors, with in-kind support from Avid, Cogstate, Albert Einstein College of Medicine, US Against Alzheimer’s disease, and Foundation for Neurologic Diseases. The companion observational Longitudinal Evaluation of Amyloid Risk and Neurodegeneration (LEARN) study is funded by the Alzheimer’s Association and GHR Foundation.

We thank the research participants, study site personnel, and all members affiliated with the HABS research team. Data used in the preparation of this article were obtained from the Harvard Aging Brain Study (HABS—P01AG036694; https://habs.mgh.harvard.edu). The HABS study was launched in 2010, funded by the National Institute on Aging, and is led by principal investigators Reisa A. Sperling MD and Keith A. Johnson MD at Massachusetts General Hospital/Harvard Medical School in Boston, MA.

Data collection and sharing for this project was funded by the Alzheimer’s Disease Neuroimaging Initiative (ADNI) (National Institutes of Health Grant U01 AG024904) and DOD ADNI (Department of Defense award number W81XWH-12-2-0012). ADNI is funded by the National Institute on Aging, the National Institute of Biomedical Imaging and Bioengineering, and through generous contributions from the following: AbbVie, Alzheimer’s Association; Alzheimer’s Drug Discovery Foundation; Araclon Biotech; BioClinica, Inc.; Biogen; Bristol-Myers Squibb Company; CereSpir, Inc.; Cogstate; Eisai Inc.; Elan Pharmaceuticals, Inc.; Eli Lilly and Company; EuroImmun; F. Hoffmann-La Roche Ltd and its affiliated company Genentech, Inc.; Fujirebio; GE Healthcare; IXICO Ltd.; Janssen Alzheimer Immunotherapy Research & Development, LLC.; Johnson & Johnson Pharmaceutical Research & Development LLC.; Lumosity; Lundbeck; Merck & Co., Inc.; Meso Scale Diagnostics, LLC.; NeuroRx Research; Neurotrack Technologies; Novartis Pharmaceuticals Corporation; Pfizer Inc.; Piramal Imaging; Servier; Takeda Pharmaceutical Company; and Transition Therapeutics. The Canadian Institutes of Health Research is providing funds to support ADNI clinical sites in Canada. Private sector contributions are facilitated by the Foundation for the National Institutes of Health (www.fnih.org). The grantee organization is the Northern California Institute for Research and Education, and the study is coordinated by the Alzheimer’s Therapeutic Research Institute at the University of Southern California. ADNI data are disseminated by the Laboratory for Neuro Imaging at the University of Southern California.

## Data Availability

Data from the A4/LEARN study is available to registered researchers through Synapse (https://www.synapse.org/Synapse:syn61250768/wiki/628717). Data from the HABS study is available to registered researchers through Synapse (https://www.synapse.org/Synapse:syn53910452/wiki/626438). Data from the ADNI study is available to registered researchers through LONI IDA (https://ida.loni.usc.edu).

## Code Availability

All scripts and resources used in this study are available via GitHub at https://github.com/JGarciaCondado/AgingBrainStudy. AgeML for training of age models is available via Github at https://github.com/compneurobilbao/ageml.

## Supplementary Tables

**Table S1.** Sample demographic by analysis type.

|  | A4/LEARN |  |  | HABS |  |  | ADNI CU |  |  | ADNI MCI |  |  |
| --- | --- | --- | --- | --- | --- | --- | --- | --- | --- | --- | --- | --- |
| Characteristic | primary<br>N = 1,635<sup>1</sup> | exploratory<br>N = 362<sup>1</sup> | p-value<sup>2</sup> | primary<br>N = 299<sup>1</sup> | exploratory<br>N = 202<sup>1</sup> | p-value<sup>2</sup> | primary<br>N = 345<sup>1</sup> | exploratory<br>N = 140<sup>1</sup> | p-value<sup>2</sup> | primary<br>N = 416<sup>1</sup> | exploratory<br>N = 82<sup>1</sup> | p-value<sup>2</sup> |
| Age | 71.62 (4.71) | 72.14 (4.85) | 0.058 | 72.30 (7.90) | 71.19 (8.29) | 0.14 | 73.24 (6.61) | 72.12 (6.55) | 0.086 | 71.93 (7.57) | 73.88 (7.08) | 0.026 |
| Sex |  |  | 0.2 |  |  | 0.6 |  |  | 0.5 |  |  | 0.2 |
| Female | 979 (59.88%) | 204 (56.35%) |  | 179 (59.87%) | 126 (62.38%) |  | 192 (55.65%) | 83 (59.29%) |  | 191 (45.91%) | 31 (37.80%) |  |
| Male | 656 (40.12%) | 158 (43.65%) |  | 120 (40.13%) | 76 (37.62%) |  | 153 (44.35%) | 57 (40.71%) |  | 225 (54.09%) | 51 (62.20%) |  |
| Education (Years) | 16.64 (2.71) | 16.28 (2.85) | 0.018 | 15.83 (2.95) | 16.07 (2.84) | 0.4 | 16.59 (2.50) | 16.52 (2.44) | 0.7 | 16.28 (2.74) | 16.32 (2.78) | >0.9 |
| APOEε4 |  |  | 0.003 |  |  | >0.9 |  |  | 0.4 |  |  | 0.4 |
| ε4- | 858 (52.48%) | 157 (43.37%) |  | 213 (71.24%) | 143 (70.79%) |  | 245 (71.01%) | 94 (67.14%) |  | 235 (56.49%) | 51 (62.20%) |  |
| ε4+ | 776 (47.46%) | 205 (56.63%) |  | 84 (28.09%) | 58 (28.71%) |  | 100 (28.99%) | 46 (32.86%) |  | 181 (43.51%) | 31 (37.80%) |  |
| Unknown | 1 (0.06%) | 0 (0.00%) |  | 2 (0.67%) | 1 (0.50%) |  | 0 (0.00%) | 0 (0.00%) |  | 0 (0.00%) | 0 (0.00%) |  |
| Aβ status |  |  | <0.001 |  |  | >0.9 |  |  | >0.9 |  |  | 0.7 |
| Aβ- | 516 (31.56%) | 33 (9.12%) |  | 225 (75.25%) | 153 (75.74%) |  | 220 (63.77%) | 89 (63.57%) |  | 191 (45.91%) | 36 (43.90%) |  |
| Characteristic | primary<br>N = 1,635<sup>1</sup> | exploratory<br>N = 362<sup>1</sup> | p-value<sup>2</sup> | primary<br>N = 299<sup>1</sup> | exploratory<br>N = 202<sup>1</sup> | p-value<sup>2</sup> | primary<br>N = 345<sup>1</sup> | exploratory<br>N = 140<sup>1</sup> | p-value<sup>2</sup> | primary<br>N = 416<sup>1</sup> | exploratory<br>N = 82<sup>1</sup> | p-value<sup>2</sup> |
| Aβ+ | 1,119<br>(68.44%) | 329 (90.88%) |  | 74 (24.75%) | 49 (24.26%) |  | 125<br>(36.23%) | 51 (36.43%) |  | 225<br>(54.09%) | 46 (56.10%) |  |
| **PACC at MRI** | 0.25 (0.58) | 0.19 (0.62) | 0.085 | -0.04 (0.67) | 0.07 (0.62) | 0.10 | 0.07 (0.58) | 0.11 (0.58) | 0.4 | -0.84 (0.81) | -0.97 (0.84) | 0.2 |
| **PACC Time from MRI**<br>**(Years)** | 0.03 (0.07) | 0.04 (0.09) | <0.001 | -0.31 (0.44) | -0.27 (0.49) | 0.12 | 0.06 (0.08) | 0.08 (0.08) | 0.042 | 0.06 (0.07) | 0.07 (0.06) | 0.010 |
| **Follow-up Time (Years)** | 5.81 (2.17) | 5.89 (1.84) | 0.4 | 8.31 (3.69) | 9.32 (3.15) | 0.004 | 6.70 (3.88) | 6.32 (3.96) | 0.3 | 5.63 (3.44) | 4.96 (3.60) | 0.062 |

<sup>1</sup>Mean (SD); n (%)
<sup>2</sup>Wilcoxon rank sum test; Pearson's Chi-squared test; Fisher's exact test

**Table S2.** Sensitivity analysis on linear mixed effects models of interaction between time from MRI and BrainAge delta for the four cohorts.

| Cohort | Quadratic vs Linear | BrainAge interaction | Adding Covariates | Placebo only |
| --- | --- | --- | --- | --- |
| A4/LEARN | **p=2e-224** | **-0.13 (p=2e-31)** | **-0.13 (p=4e-27)** | **-0.16 (p=4e-15)** |
| HABS | **p=1.1e-29** | **-0.19 (p=2e-8)** | **-0.16 (p=2.1e-8)** | N/A |
| ADNI CU | **p=1.6e-34** | -0.06 (p=0.13) | **-0.08 (p=0.04)** | N/A |
| ADNI MCI | **p=1.2e-17** | **-0.31 (p=4e-16)** | **-0.29 (p=9.4e-15)** | N/A |

**Table S3.** Individual interaction terms between time from MRI and BrainAge delta or biomarkers for each of the biomarkers in each of the cohorts.

| Cohot | Aβ-PET | | | pTau<sub>217</sub> | | | Tau-PET | | |
| --- | --- | --- | --- | --- | --- | --- | --- | --- | --- |
| | BrainAge delta | Aβ-PET | BrainAge delta x Aβ-PET | BrainAge delta | pTau<sub>217</sub> | BrainAge delta x pTau<sub>217</sub> | BrainAge delta | Tau-PET | BrainAge delta x Tau-PET |
| A4/LEARN | -0.11 (p=0.24) | **-0.17 (p=8e-26)** | **-0.08 (p=9e-15)** | -0.14 (p=0.69) | **-0.15 (p=2.0e-9)** | **-0.08 (p=4e-5)** | -0.08 (p=0.11) | **-0.27 (p=7e-29)** | **-0.04 (p=0.02)** |
| HABS | -0.12 (p=0.69) | **-0.25 (p=1.2e-18)** | **-0.14 (p=7.7e-8)** | -0.11 (p=0.81) | **-0.25 (p=1.2e-15)** | **-0.1 (p=7.9e-5)** | **-0.08 (p=8.4e-9)** | **-0.27 (p=3.8e-22)** | **-0.16 (p=2.1e-10)** |
| ADNI CU | -0.04 (p=0.97) | **-0.25 (p=2.2e-12)** | **-0.08 (p=0.007)** | -0.15 (p=0.30) | **-0.33 (p=2.3e-6)** | -0.031 (p=0.66) | -0.05 (p=0.70) | **-0.34 (p=6.2e-8)** | 0.02 (p=0.78) |
| ADNI MCI | **-0.2 (p=0.01)** | **-0.33 (p=8.3e-13)** | **-0.11 (p=0.003)** | -0.10 (p=0.11) | **-0.29 (p=0.02)** | **-0.26 (p=0.003)** | -0.07 (p=0.16) | **-0.35 (p=0.005)** | -0.19 (p=0.1) |

**Table S4.** Sensitivity analysis of interaction term of BrainAge delta and biomarkers in linear mixed effects models when including, sex, age at MRI, education, Aβ status and *APOE*ε4 carriers.

| Cohort | Aβ-PET | pTau<sub>217</sub> | Tau-PET |
| --- | --- | --- | --- |
| A4/LEARN | **-0.08 (p=7.2e-14)** | **-0.08 (p=2.8e-5)** | **-0.04 (p=0.016)** |
| HABS | **-0.14 (p=1.7e-8)** | **-0.10 (p=6.2e-5)** | **-0.15 (p=1.3e-9)** |
| ADNI CU | **-0.09 (p=0.002)** | -0.05 (p=0.46) | 0.03 (p=0.72) |
| ADNI MCI | **-0.11 (p=0.01)** | **-0.29 (p=0.003)** | -0.19 (p=0.12) |

**Table S5.** Sensitivity analysis of interaction term of BrainAge delta and biomarkers in linear mixed effects models when including, sex, age at MRI, education, Aβ status and *APOE*ε4 carriers for biomarker measurements taken within a year of the MRI.

| Cohort | Aβ-PET | pTau<sub>217</sub> | Tau-PET |
| --- | --- | --- | --- |
| A4/LEARN | **-0.08 (p=1.3e-13)** | **-0.08 (p=2.8e-5)** | **-0.04 (p=0.017)** |
| HABS | **-0.14 (p=1.6e-8)** | **-0.10 (p=0.0002)** | **-0.09 (p=0.002)** |
| ADNI CU | -0.08 (p=0.05) | 0.29 (p=0.16) | 0.10 (p=0.09) |
| ADNI MCI | **-0.13 (p=0.0002)** | -0.15 (p=0.44) | -0.01 (p=0.89) |

## Supplementary Figures

**Figure S1.**
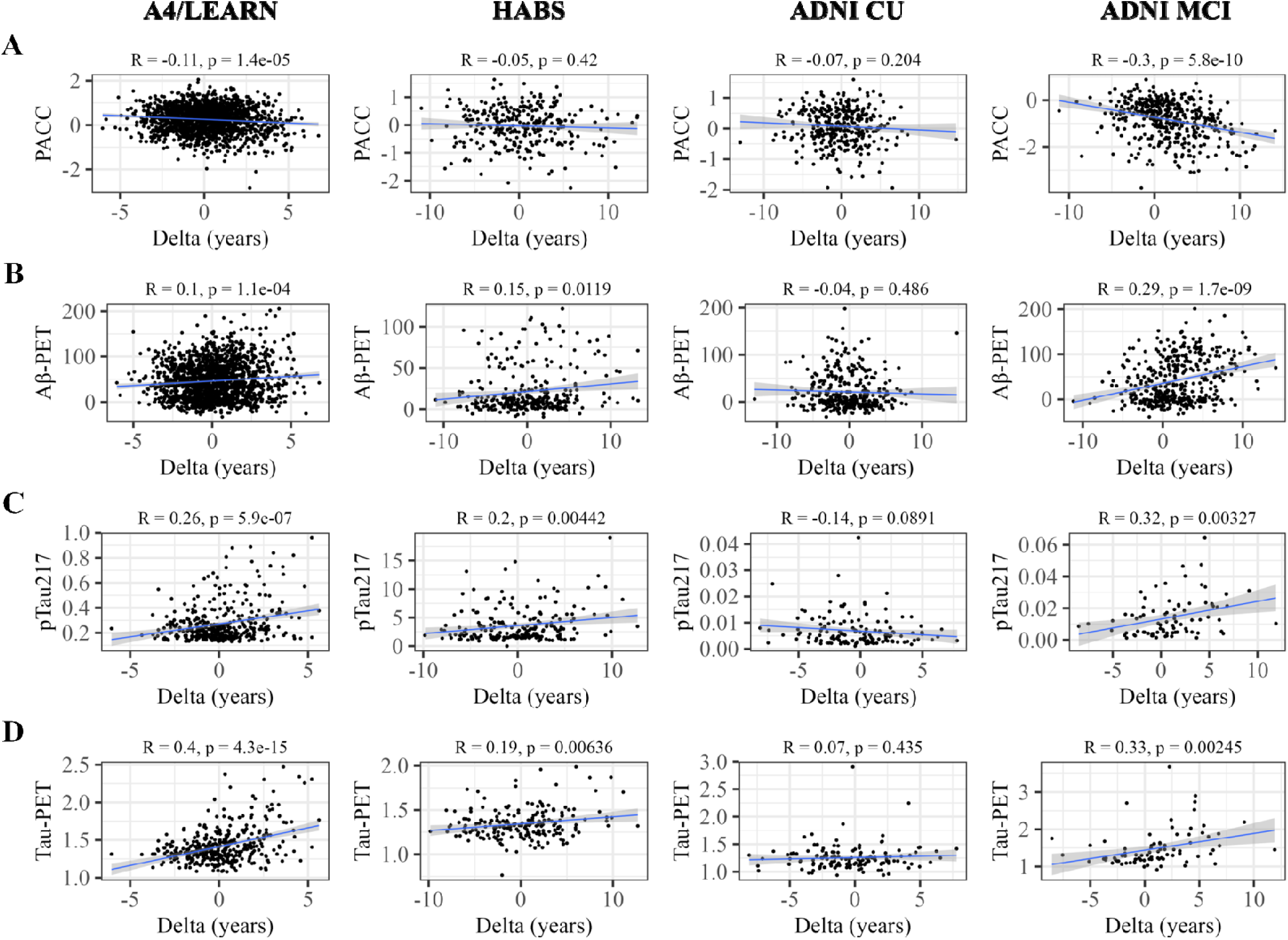
Correlation of BrainAge delta with clinical and biomarker measures closest in time to MRI. (**A)** Correlation with PACC. **(B)** Correlation with Aβ-PET. **(C)** Correlation with plasma pTau_217_. **(D)** Correlation with Tau-PET. Each column shows results for each cohort. Ranges for the y-axis vary for each biomarker across cohorts except for PACC that has been homogenized, and Aβ-PET is in centiloids. Each cohort had different measurement types of Aβ-PET, pTau_217_ and Tau-PET. The x-axis BrainAge delta varies as different models were trained for each cohort. ADNI MCI displays higher BrainAge deltas than ADNI CU.

**Figure S2.**
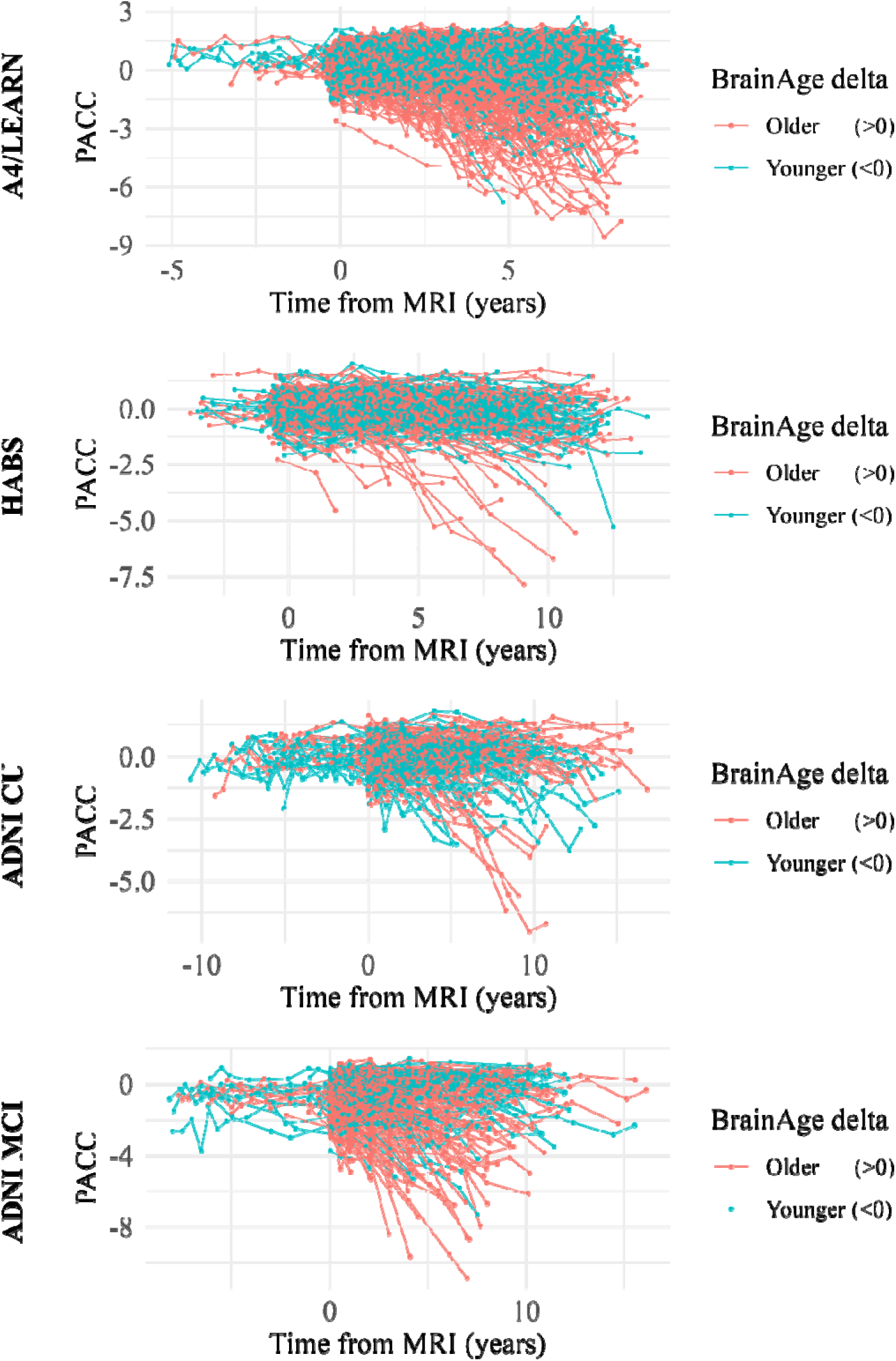
Individual longitudinal cognitive trajectories for each of the four cohorts.

**Figure S4.**
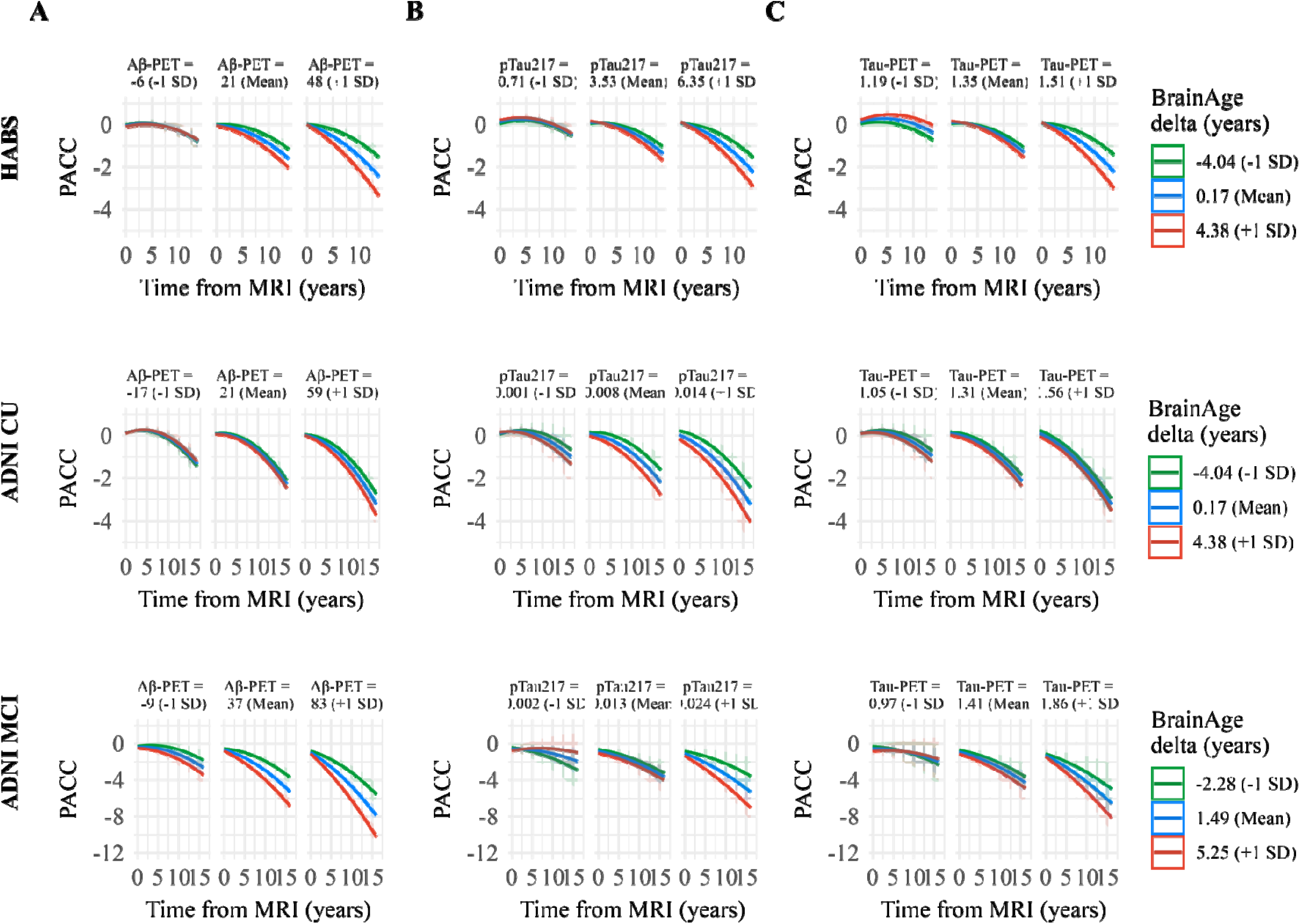
Linear mixed effects models for longitudinal cognitive trajectories of clinical trials. **(A)** Linear mixed effects model for ADNI MCI using CDRSUM of boxes for (1) only Aβ positive participants and (2) only Aβ positive participants with BrainAge delta above the median. **(B)** Linear mixed effects models for A4 placebo group using PACC.

**Figure S5.**
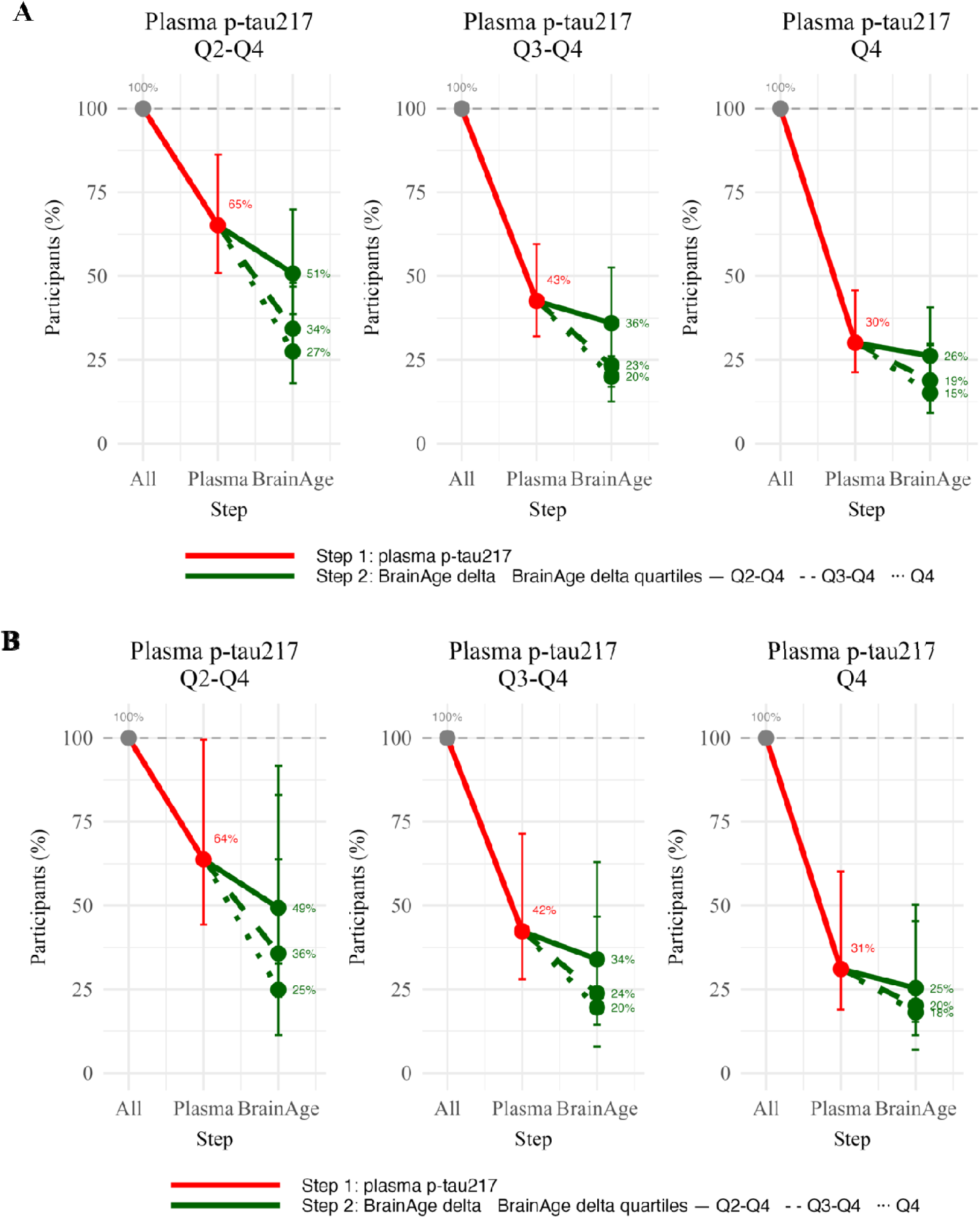
Screening for pTau217 and BrainAge on A4 and A4 placebo cohorts. The obtained sample size reduction using sample selection based on different percentiles (75^th^ [Q2-Q4], 50^th^ [Q3-Q4] and 25^th^ [Q4]) of baseline plasma pTau_217_ levels in step 1 followed by selection based on the same percentiles (75th, 50th and 25th) of the BrainAge measurement in step 2 with PACC as the primary endpoint. Error bars represent the 95% CI around the mean derived from linear effects models. Red lines represent step 1 with plasma p-tau217, and green lines represent step 2 with BrainAge. Different line styles represent different quartiles of BrainAge from those participants already selected from step 1. Calculations in assume 80% power to detect a 30% change in PACC in a 4-year trial. **(A)** Using data from only the A4 study which includes participants with elevated Aβ-PET **(B)** Using data from only the A4 placebo group.

## Notes

### Competing Interest Statement

The authors have declared no competing interest.

### Author Declarations

We conducted the procedures for this study under the ethical guidelines stipulated by the Massachusetts General Brigham Human Research Committee, which is the relevant Institutional Review Board. The study was carried out by the guidelines of the Declaration of Helsinki. Written informed consent was obtained from all participants. The data used in this study is from individual-level data that has been de-identified prior to its use in this study.

### Summary of Updates

The discussion has been updated to reflect ongoing discussions on the utility of BrainAge for clinical trials. Figure 3 and 4 were updated with clear meta-analysis tables.

## References

1. Franke, K., Ziegler, G., Klöppel, S. & Gaser, C. Estimating the age of healthy subjects from T1-weighted MRI scans using kernel methods: Exploring the influence of various parameters. NeuroImage 50, 883–892 (2010).

2. Gaser, C. et al. BrainAGE in Mild Cognitive Impaired Patients: Predicting the Conversion to Alzheimer’s Disease. PLoS ONE 8, e67346 (2013).

3. Van Der Flier, W. M. & Scheltens, P. The ATN Framework—Moving Preclinical Alzheimer Disease to Clinical Relevance. JAMA Neurol 79, 968 (2022).

4. Boyle, P. A. et al. To what degree is late life cognitive decline driven by age-related neuropathologies? Brain 144, 2166–2175 (2021).

5. Gonzales, M. M. et al. Biological aging processes underlying cognitive decline and neurodegenerative disease. Journal of Clinical Investigation 132, e158453 (2022).

6. Abughofah, Y., et al. Association between BrainAGE and Alzheimer’s disease biomarkers. Alz & Dem Diag Ass & Dis Mo 17, e70094 (2025).

7. Löwe, L. C., Gaser, C., Franke, K., & for the Alzheimer’s Disease Neuroimaging Initiative. The Effect of the APOE Genotype on Individual BrainAGE in Normal Aging, Mild Cognitive Impairment, and Alzheimer’s Disease. PLoS ONE 11, e0157514 (2016).

8. Cumplido-Mayoral, I. et al. Biological brain age prediction using machine learning on structural neuroimaging data: Multi-cohort validation against biomarkers of Alzheimer’s disease and neurodegeneration stratified by sex. eLife 12, e81067 (2023).

9. Boyle, R. et al. Brain-predicted age difference score is related to specific cognitive functions: a multi-site replication analysis. Brain Imaging and Behavior 15, 327–345 (2021).

10. Beheshti, I., Maikusa, N. & Matsuda, H. The association between “Brain-Age Score” BAS and traditional neuropsychological screening tools in Alzheimer’s disease. Brain and Behavior 8, e01020 (2018).

11. Rissman, R. A. et al. Plasma Aβ42/Aβ40 and phospho-tau217 concentration ratios increase the accuracy of amyloid PET classification in preclinical Alzheimer’s disease. Alzheimer’s & Dementia 20, 1214–1224 (2024).

12. Barthélemy, N. R. et al. Highly accurate blood test for Alzheimer’s disease is similar or superior to clinical cerebrospinal fluid tests. Nat Med 30, 1085–1095 (2024).

13. Van Dyck, C. H. et al. Lecanemab in Early Alzheimer’s Disease. N Engl J Med 388, 9–21 (2023).

14. Chen, C.-L. et al. Validation of neuroimaging-based brain age gap as a mediator between modifiable risk factors and cognition. Neurobiology of Aging 114, 61–72 (2022).

15. Cumplido-Mayoral, I. et al. The mediating role of neuroimaging-derived biological brain age in the association between risk factors for dementia and cognitive decline in middle-aged and older individuals without cognitive impairment: a cohort study. The Lancet Healthy Longevity 5, e276–e286 (2024).

16. Cumplido-Mayoral, I. et al. Neuroimaging-derived biological brain age and its associations with glial reactivity and synaptic dysfunction cerebrospinal fluid biomarkers. Mol Psychiatry 10.1038/s41380-025-02961-x (2025) doi:10.1038/s41380-025-02961-x.

17. Howe, M. D. et al. Clinical application of plasma P-tau217 to assess eligibility for amyloid-lowering immunotherapy in memory clinic patients with early Alzheimer’s disease. Alz Res Therapy 16, 154 (2024).

18. Ossenkoppele, R. et al. Plasma p-tau217 and tau-PET predict future cognitive decline among cognitively unimpaired individuals: implications for clinical trials. Nat Aging 5, 883–896 (2025).

19. Sims, J. R. et al. Donanemab in Early Symptomatic Alzheimer Disease: The TRAILBLAZER-ALZ 2 Randomized Clinical Trial. JAMA 330, 512 (2023).

20. Sperling, R. A. et al. Amyloid-related imaging abnormalities in amyloid-modifying therapeutic trials: Recommendations from the Alzheimer’s Association Research Roundtable Workgroup. Alzheimer’s & Dementia 7, 367–385 (2011).

21. Schulz, M.-A., Siegel, N. T. & Ritter, K. Brain-age models with lower age prediction accuracy have higher sensitivity for disease detection. Preprint at 10.1101/2024.03.28.587212 (2024).

22. Garcia Condado, J., et al. Aging as an active player in Alzheimer’s disease classification: Insights from feature selection in BrainAge models. NeuroImage 322, 121548 (2025).

23. Sperling, R. A. et al. The A4 Study: Stopping AD Before Symptoms Begin? Sci. Transl. Med. 6, (2014).

24. Dagley, A. et al. Harvard Aging Brain Study: Dataset and accessibility. NeuroImage 144, 255–258 (2017).

25. Aisen, P. S. et al. Clinical core of the Alzheimer’s disease neuroimaging initiative: Progress and plans. Alzheimer’s & Dementia 6, 239–246 (2010).

26. Duff, K. & on behalf of the ADNI investigators. Amnestic MCI in ADNI: Maybe Not Enough Memory Impairment? Neurology 97, 595–596 (2021).

27. Fischl, B. FreeSurfer. NeuroImage 62, 774–781 (2012).

28. Sabuncu, M. R. The Dynamics of Cortical and Hippocampal Atrophy in Alzheimer Disease. Arch Neurol 68, 1040 (2011).

29. Poulin, S. P., Dautoff, R., Morris, J. C., Barrett, L. F. & Dickerson, B. C. Amygdala atrophy is prominent in early Alzheimer’s disease and relates to symptom severity. Psychiatry Research: Neuroimaging 194, 7–13 (2011).

30. Bakkour, A., Morris, J. C. & Dickerson, B. C. The cortical signature of prodromal AD: Regional thinning predicts mild AD dementia. Neurology 72, 1048–1055 (2009).

31. Landau, S. M. et al. Amyloid deposition, hypometabolism, and longitudinal cognitive decline. Annals of Neurology 72, 578–586 (2012).

32. Sperling, R. A. et al. Association of Factors With Elevated Amyloid Burden in Clinically Normal Older Individuals. JAMA Neurol 77, 735 (2020).

33. Klunk, W. E. et al. The Centiloid Project: Standardizing quantitative amyloid plaque estimation by PET. Alzheimer’s & Dementia 11, 1 (2015).

34. Rentz, D. M. et al. Characterizing Clinical Progression in Cognitively Unimpaired Older Individuals with Brain Amyloid: Results from the A4 Study. The Journal of Prevention of Alzheimer’s Disease 11, 814–822 (2024).

35. Farrell, M. E. et al. Defining the Lowest Threshold for Amyloid-PET to Predict Future Cognitive Decline and Amyloid Accumulation. Neurology 96, (2021).

36. for the Alzheimer’s Disease Neuroimaging Initiative et al. Validation of amyloid PET positivity thresholds in centiloids: a multisite PET study approach. Alz Res Therapy 13, 99 (2021).

37. Johnson, K. A. et al. Tau positron emission tomographic imaging in aging and early A lzheimer disease. Annals of Neurology 79, 110–119 (2016).

38. Young, C. B. et al. Divergent Cortical Tau Positron Emission Tomography Patterns Among Patients With Preclinical Alzheimer Disease. JAMA Neurol 79, 592 (2022).

39. Maass, A. et al. Comparison of multiple tau-PET measures as biomarkers in aging and Alzheimer’s disease. NeuroImage 157, 448–463 (2017).

40. the Alzheimer’s Disease Neuroimaging Initiative et al. Four distinct trajectories of tau deposition identified in Alzheimer’s disease. Nat Med 27, 871–881 (2021).

41. Donohue, M. C. et al. The Preclinical Alzheimer Cognitive Composite: Measuring Amyloid-Related Decline. JAMA Neurol 71, 961 (2014).

42. Papp, K. V., Rentz, D. M., Orlovsky, I., Sperling, R. A. & Mormino, E. C. Optimizing the preclinical Alzheimer’s cognitive composite with semantic processing: The PACC5. A&D Transl Res & Clin Interv 3, 668–677 (2017).

43. Mormino, E. C. et al. Early and late change on the preclinical Alzheimer’s cognitive composite in clinically normal older individuals with elevated amyloid β. Alzheimer’s & Dementia 13, 1004–1012 (2017).

44. Hampton, O. L. et al. Harmonizing the preclinical Alzheimer cognitive composite for multicohort studies. Neuropsychology 37, 436–449 (2023).

45. Sperling, R. A. et al. Trial of Solanezumab in Preclinical Alzheimer’s Disease. N Engl J Med 389, 1096–1107 (2023).

46. Rissman, R. A. et al. Longitudinal Phospho-tau217 Predicts Amyloid Positron Emission Tomography in Asymptomatic Alzheimer’s Disease. The Journal of Prevention of Alzheimer’s Disease 11, 823–830 (2024).

47. Hu, Y. et al. Assessment of a Plasma Amyloid Probability Score to Estimate Amyloid Positron Emission Tomography Findings Among Adults With Cognitive Impairment. JAMA Netw Open 5, e228392 (2022).

48. Leitão, M. J. et al. Clinical validation of the Lumipulse G cerebrospinal fluid assays for routine diagnosis of Alzheimer’s disease. Alz Res Therapy 11, 91 (2019).

49. Garcia Condado, J., Elorriaga, I. T., Cortes, J. M. & Erramuzpe, A. AgeML: Age modeling with Machine Learning. IEEE J. Biomed. Health Inform. 1–11 (2025) doi:10.1109/JBHI.2025.3531017.

50. Birkenbihl, C. et al. Rethinking the residual approach: leveraging statistical learning to operationalize cognitive resilience in Alzheimer’s disease. Brain Inf. 12, 3 (2025).

51. Buckley, R. F. et al. Sex, amyloid, and *APOE* ε4 and risk of cognitive decline in preclinical Alzheimer’s disease: Findings from three well-characterized cohorts. Alzheimer’s & Dementia 14, 1193–1203 (2018).

52. Sperling, R. A. et al. Amyloid and Tau Prediction of Cognitive and Functional Decline in Unimpaired Older Individuals: Longitudinal Data from the A4 and LEARN Studies. The Journal of Prevention of Alzheimer’s Disease 11, 802–813 (2024).

53. Birkenbihl, C. et al. Artificial intelligence-based clustering and characterization of Parkinson’s disease trajectories. Sci Rep 13, 2897 (2023).

54. Petersen, R. C. MCI Criteria in ADNI: Meeting Biological Expectations. Neurology 97, 597–599 (2021).

55. Morris, J. C. The Clinical Dementia Rating (CDR): Current version and scoring rules. Neurology 43, 2412 (1993).

56. Ard, M. C. & Edland, S. D. Power Calculations for Clinical Trials in Alzheimer’s Disease. JAD 26, 369–377 (2011).

57. Iddi, S. & Donohue, M. C. Power and Sample Size for Longitudinal Models in R – The longpower Package and Shiny App. The R Journal 14, 264–282 (2022).

58. Birkenbihl, C., De Jong, J., Yalchyk, I. & Fröhlich, H. Deep learning-based patient stratification for prognostic enrichment of clinical dementia trials. Brain Communications 6, fcae445 (2024).

59. Jönsson, L. et al. The affordability of lecanemab, an amyloid-targeting therapy for Alzheimer’s disease: an EADC-EC viewpoint. The Lancet Regional Health - Europe 29, 100657 (2023).

60. Ross, E. L., Weinberg, M. S. & Arnold, S. E. Cost-effectiveness of Aducanumab and Donanemab for Early Alzheimer Disease in the US. JAMA Neurol 79, 478 (2022).

